# Pretrained transformers applied to clinical studies improve predictions of treatment efficacy and associated biomarkers

**DOI:** 10.1101/2023.09.12.23295357

**Authors:** Gustavo Arango-Argoty, Elly Kipkogei, Ross Stewart, Arijit Patra, Ioannis Kagiampakis, Etai Jacob

## Abstract

Cancer treatment has made significant advancements in recent decades, leading to improved outcomes and quality of life for many patients. Despite the array of available therapies, including targeted, hormone, and checkpoint blockade immunotherapy, many patients experience treatment failure or eventual resistance. Attempts to predict the efficacy of therapies, particularly immuno-oncology therapies, have suffered from limited accuracy and difficulties in identifying molecular and other determinants of response. Improving treatment prediction alone is insufficient to create clinically meaningful research tools; additional prerequisites for this goal involve accommodating small data sets, effectively handling sparse features, integrating diverse clinical data, addressing missing measurements, ensuring interpretability, and extracting valuable biological insights for both clinical context and further research. Multimodal deep-learning models offer a promising avenue to surmount these challenges by leveraging their capacity and flexibility to learn from expansive and varied clinical and molecular data sets. Similar to their application in natural language and other domains, deep-learning models can uncover complex relationships within data that are pertinent to survival and treatment response. In this study, we introduce an explainable transformer-based deep-learning framework that addresses these challenges. This framework yields predictions of survival outcomes, as quantified by concordance index, that surpass the performance of state-of-the-art methods such as Cox proportional hazards, survival random forest, and tumor mutation burden, across diverse independent data sets. We developed the clinical transformer, a deep neural-network survival prediction framework that capitalizes on the flexibility of the deep-learning model, including training strategies like gradual and transfer learning, to maximize the use of available data to enhance survival predictions and generate actionable biological insights. Finally, we illustrate the future potential of the clinical transformer’s generative capability in early-stage clinical studies. By perturbing molecular features associated with immune checkpoint inhibition treatment in immunotherapy-naive patient profiles, we identified a subset of patients who may benefit from immunotherapy. These findings were subsequently validated across three independent immunotherapy treatment cohorts. We anticipate that this research will empower the scientific community to further harness data for the benefit of patients.

---

The emergence of immunotherapy based on anti–programmed cell death 1 (PD-1) and its ligand (PD-L1) has transformed the treatment of cancer over the last decade. However, despite dramatic and durable responses, in most patients the disease progresses, and in some it fails to respond initially. Anti–PD-1, anti–PD-L1, and other checkpoint blockade approaches do not directly target the tumor but instead recruit the patient’s immune system to fight the disease. Due to this indirect mechanism of action, the drivers of response to therapy are more complicated than those of, for example, a targeted inhibitor of an oncogene and can be affected by the patient’s physical condition and immune system fitness, as well as the tumor’s underlying biology. Despite advancements in precision medicine, biomarker discovery,^1, 2^ and machine learning–based modeling,^3–5^ our ability to identify patients who will respond to treatment is still limited, as is as our means to understand the mechanism behind resistance.^6, 7^ At present, only three predictive biomarkers have received approval from the U.S. Food and Drug Administration (FDA) for use as companion diagnostics for immunotherapy. These biomarkers are the assessment of microsatellite instability (MSI), tumor PD-L1 expression by immunohistochemistry, and the measurement of tumor mutation burden (TMB).^8–10^

In recent years, transformers^11^ have been extensively used in state-of-the-art applications for natural-language modeling. Examples include chatGPT,^12, 13^ image processing (e.g., Dino,^14^ ViT),{Liu, 2023’; online ahead of print #24;Chen, 2022 #2} protein structure prediction (e.g., alphaFold),^15, 16^ and de novo protein sequence generation.^17, 18^ One of the key features of transformers in all these applications is self-attention, a mechanism designed to draw dependencies among all features.^19^ Self-attention is especially important for language processing tasks, in which the meaning of a word can change based on its context within a sentence. As an analogy, in precision medicine, where one wishes to predict the best treatment for a patient, a biomarker may have limited meaning if it is measured independently of other clinical or molecular features. Transformers could be used to weigh the importance of various biomarkers in a given patient’s disease in the context of all available clinical and molecular measurements and to dynamically adjust their influence on the output, such as response to treatment. Therefore, the potential of an attention mechanism to capture the complexities in patients’ characteristics and response, as successfully captured in applications such as natural-language processing and understanding, could improve our ability to predict patient outcomes.

Improving the prediction of treatment efficacy or survival by itself, however, is insufficient to establish an effective research tool that can offer informative guidance and insights for clinical decision-making. Additional requirements must be met to build models that will potentially have a translational impact. First, models must be compatible with relatively small data sets, such as those in clinical studies (e.g., patient cohorts in phase 1–3 clinical trials typically include 100 or fewer to 1000 patients), in contrast to training data in other areas of research where deep learning is applied (e.g., imaging, natural-language processing). Second, models must be able to adeptly manage sparse features, such as functional mutation events from genomic profiling, that are typically infrequent within a patient population. Third, predictive models are required to incorporate data from multiple modalities available in clinical studies and real-world data, including, for example, features derived from DNA and RNA sequencing from tumor or peripheral blood, clinical and temporal measurements, proteomics, methylomics, and more. In addition, some measurements could be missing in more than one part of a patient cohort (e.g., due to unsuccessful assays or biopsies or limited measurements). In addition to these challenges, biological and clinical data are noisy, variable, and inconsistent (e.g., batch effects or data generated by different assays or laboratories). Therefore, to maximize the use of artificial intelligence (AI) machinery for clinical practice, it must have the capability to effectively integrate high-dimensional and diverse data, combine patient populations, and handle missing data. Fourth, although deep-learning models are often seen as “black boxes” (i.e, a computational model or algorithm that provides predictions or decisions without offering insight into how those predictions were derived), in order to translate models to the clinical domain it is essential to transition to interpretable or linear models, or even simple rule-based approaches like decision trees. Therefore, explainable AI is necessary to allow researchers to examine whether the deep-learning prediction model aligns with existing knowledge, increases confidence in survival predictions, and relates the results to the relevant clinical context (e.g., to associate better survival predictions with features that support a certain type of treatment). Finally, the ability to extract biological insights and understand disease biology in the relevant clinical context from prediction models is vital for effectively informing further research. This requirement includes providing a mechanistic understanding of drug effects, identifying potential molecular targets, and elucidating mechanisms of resistance to treatment.

To address these needs, we created the clinical transformer, a deep neural-network survival prediction framework based on transformers. This framework effectively handles relatively small data sets by incorporating a transfer learning mechanism. It leverages large data sets (e.g., The Cancer Genome Atlas [TCGA] and the Genomics Evidence Neoplasia Information Exchange [GENIE]) to build a foundation model that is then fine-tuned on specific smaller data sets (e.g., a patient cohort from a clinical study). The clinical transformer exhibited flexibility in handling diverse data sets with various feature types and levels of sparsity, as well as the ability to combine patient populations. Owing to the attention mechanism in the clinical transformer architecture, it was able to capture complex, nonlinear relationships among multiple molecular, clinical, and demographic feature modalities of patients. We trained and tested our model with seven pan-cancer data sets comprising more than 140,000 patients from immuno-oncologic (IO), targeted, and chemotherapy treatments. Table 1 lists the data sets used in this study in addition to the various indications (cancer-specific or pan-cancer) and the use of the data (described in the “data usage” column in the table for pretraining, survival, or independent validation). When we then compared the model’s performance against those of state-of-the-art methods used for survival prediction, we found that our model consistently outperformed these methods, including Cox proportional hazards (PH) and random survival forest. Here we describe how the internal representations learned by the fully trained model could be used for specialized tasks, such as prediction of response to immunotherapy, in small real-world data sets or early-stage clinical trials. Finally, we show how the clinical transformer’s explainability and perturbation modules could be used to improve clinical interpretations of outcome and response predictions by allowing researchers to identify the molecular and clinical features that lead to specific outcomes in individual patients.

**Table 1.** Data sets used in this study.

| **Data set (reference)** | **Cancer type** | **Description** | **Data usage** | **No. of patients** | **Treatment** | **No. of features** | **Features** |
| --- | --- | --- | --- | --- | --- | --- | --- |
| Chowell et al. 2021<sup>3</sup> | Pan-cancer | Patients treated with anti-PD-1/PD-L1 and combo in multiple cancer types | Survival modeling (training and testing), transfer learning | 1479 | IO | 17 | Data set consisting of 17 features (e.g., TMB, HLA-LOH, MSI, albumin, NLR) |
| MYSTIC trial (NCT02453282) | NSCLC | Stage IV NSCLC treatment-naïve anti-PD-L1 and anti-PD-L1 + anti-CTLA-4 | Survival modeling (training and testing) | 325 | IO | 15 | Data set consisting of 15 features (e.g., TMB, HLA-LOH, MSI, albumin, NLR) in addition to FMI mutation calls, in addition to a subset of 150 with HLA germline typing |
| OAK trial (NCT02008227) | NSCLC | Stage IV NSCLC patients treated with anti-PD-1/PD-L1 after failure of chemotherapy | Survival modeling (training and testing) | 396 | IO | 418 | ctDNA-derived mutation calls |
| Samstein et al. 2019<sup>25</sup> | Pan-cancer | Response to IO treatment (anti-PD-1/PD-L1 and combo) in pan-cancer setting | Survival modeling (training and testing) | 1610 | IO | 474 | Tissue MSK-IMPACT calls from 474 genes in addition to demographic features |
| Thorsson et al. 2018<sup>24</sup> | Pan-cancer | Study on TCGA data that derives features associated with the tumor micro environment in a pan-cancer setting. | Survival modeling (training and testing) | 6012 | TCGA | 49 | A total of 49 derived features from RNA sequencing data that profile the TME |
| Bagaev et al. 2021<sup>56</sup> | Pan-cancer | TME subtypes derived from TCGA data | Pretraining and survival modeling (training and testing) for SKCM | 11070 | TCGA | 29 | A total of 29 RNA signatures obtained from this study |
| Van Allen et al.<sup>50</sup> – DFCI metastatic melanoma | Melanoma | Metastatic melanoma data set from DFCI; anti-CTLA-4 monotherapy | Validation data set | 110/40 | IO | 36,859 | Whole exomes from pretreatment melanoma tumor biopsies and matching germline tissue samples from 110 patients |
| AACR project GENIE (Pugh et al.<sup>21</sup>) | Pan-cancer | Publicly accessible international cancer registry of real-world data assembled through data sharing among 19 of the leading cancer centers in the world | Transfer learning | 134,626 | Not available | 2290 | Mutation and copy number calls from multiple panels and centers spanning 2290 genes in addition to demographic features |
| MSK MIND (Vanguri et al.<sup>4</sup>) | Lung | Initiative to accelerate research and discovery through advanced analytics | Validation data set | 247 | IO | 1043 | Cohort of 247 patients with advanced NSCLC with multimodal baseline data; only molecular data used |
| Miao et al. 2018<sup>49</sup> | Pan-cancer | WES of 249 tumors from patients with clinically annotated outcomes to immune checkpoint therapy | Validation data set | 249 | IO | >10,000 | WES and demographic features |
| Riaz et., al 2017<sup>58</sup> | Melanoma | Whole transcriptome from patients who progressed on ipilimumab or were ipilimumab naive, | Validation data set | 26 | IO | 29 | Extracted 29 signatures defined by the Bagaev et al.<sup>56</sup> signatures using whole transcriptome data |

| **Data set**<br>**(reference)** | **Cancer**<br>**type** | **Description** | **Data usage** | **No. of**<br>**patients** | **Treatment** | **No. of**<br>**features** | **Features** |
| --- | --- | --- | --- | --- | --- | --- | --- |
|  |  | before and<br>after nivolumab |  |  |  |  |  |
| Liu et al. 2019<sup>57</sup> | Melanoma | Whole transcriptome for<br>patients treated with<br>anti-PD-1 therapy | Validation data set | 42 | IO | 29 | Extracted 29 signatures defined<br>by the Bagaev et al.<sup>56</sup> signatures<br>using whole transcriptome data |

ctDNA, circulating tumor DNA; DFCI, Dana Farber Cancer Institute; FMI, Foundation Medicine; WES, whole-exome sequencing.

## RESULTS

### The clinical transformer framework

The clinical transformer framework is shown in Fig. 1a and includes the following elements: input agnostic modeling and integration to enable prediction using multiple molecular (e.g., omics), clinical, and demographic modalities and their integration in a single system, including handling sparse features, missing data, and different annotation hierarchies (e.g., gene vs. protein name) (Fig. 1b); self-supervision and transfer learning to enable analysis on relatively small data sets, as in clinical studies (Fig. 1c); an interpretability module to enable executions of clinical applications that require high confidence and understanding of the prediction model, including the capability to suggest biological insights (Fig. 1a); and a generative model to enable synthetic generation of populations of patients with distinct characteristics that may not currently exist in our data set. This was achieved by perturbing the molecular or clinical features of real patients and predicting the new virtual response after these modifications. This generative AI capability can enable researchers to explore scenarios that contribute to resistance or enhance response across a diverse array of patient populations (Fig. 1a).

**Fig. 1.**
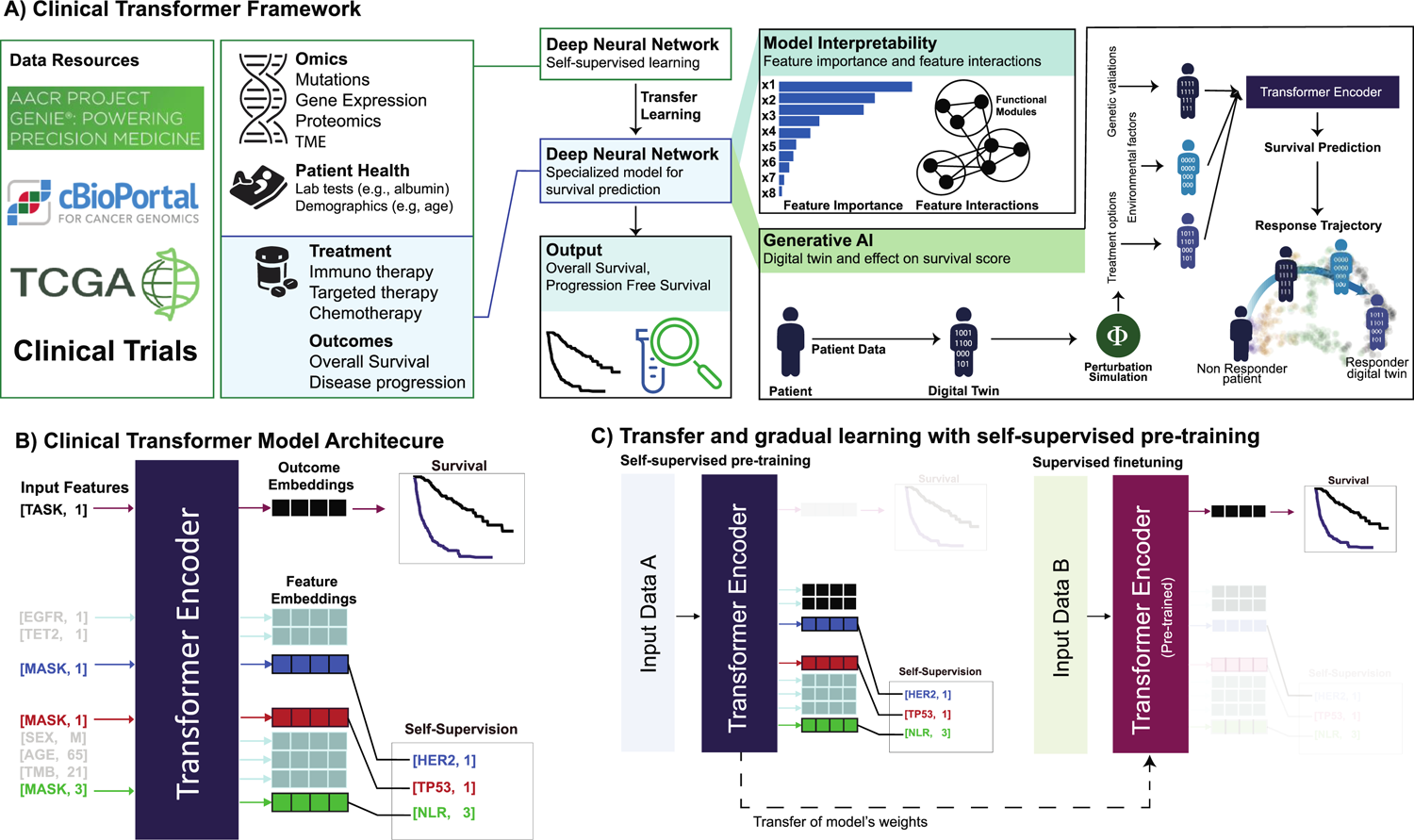
Overview of the clinical transformer framework. **a**, Framework capabilities and analysis overview of the clinical transformer to process data to insights. Model interpretability is extracted by using the output embeddings to generate functional modules (group or input features) that are associated with the outcome. The clinical transformer is shown as a generative model to recreate a patient’s trajectory of response, using patient embeddings with a perturbation-based approach. Embeddings can also generate synthetic data restricted by certain conditions. **b**, Input data are represented by [Key, Value] pairs, where the key is the feature name and the value corresponds to the numerical score of the feature (e.g., [Age, 20] represents a patient with an age of 20 years). Feature names and values are embedded and fed to a transformer encoder architecture without positional encoding. The special input token [TASK, 1] is added in front of every input sample, and the output of this token is used to predict patient survival or classification outcomes. The special token [MASK] is used for performing the pretraining stage, in which the model is asked to predict the masked token names (corresponding to the masked features). **c**, The clinical transformer trained in self-supervised mode uses data set A to train over a masked-prediction task in which input features are randomly ignored and used as labels. Thus, the objective of the model is to predict the feature name of the ignored input features. After pretraining, the weights of the model can be used to fine-tune into a specialized task such as responder prediction or survival analysis. When input data A are different from input data B, it refers to transfer learning, whereas if the same data are used for both tasks (data A = data B), it is called gradual learning, as the model first learns about the data in an unsupervised way and then specializes on a specific task over the same data set (e.g., survival prediction).

### Learning strategies

Our modeling framework is composed of three learning strategies: (1) direct learning, in which the model is trained from scratch to perform a given task, such as survival or response prediction; (2) gradual learning, in which, using the same input data for survival prediction, the model is first trained with self-supervised learning for masked feature prediction, similar to the strategy used to train large language models such as BERT^20^ (Fig. 1c), and is then fine-tuned on a specific task (e.g., patient response or survival prediction); and (3) transfer learning, in which a model is pretrained on large amounts of data, using self-supervision for masked feature prediction, and the weights are then used to initialize other models that are fine tuned to predict survival or patient response. The pretraining step (i.e., gradual learning and/or transfer learning) allows the model to recognize nonspecific biological and clinical patterns and relationships in the data. This step provides a more informative starting point than a random initialization and enables a new model to be further refined on another or a similar data set, focusing on patient outcome as a target function (Fig. 1c).

### Data sets used in training, validation, and testing

In total, 12 data sets from clinical trials and real-world data (pan- and cancer specific), comprising a total of 150,070 patients, were used in the framework for multiple tasks, including the three strategies mentioned above: direct, gradual, and transfer learning (Table 1). Data sets were used independently to predict patient response to treatment in direct and gradual learning modes, as well as by combining data sets with similar input features to evaluate the impact of transfer learning. All data sets were used to predict patient response to treatment with the survival outcomes, except for the GENIE project’s data, which was used in the self-supervision mode for transfer learning. These data comprised 134,626 patients^21^ and were used to build a model based on the gradual learning strategy mentioned above, using mutations and demographics data as input (Table 1). Instances of this general-purpose model were further used in multiple independent tasks to predict patient survival after fine-tuning based on relatively small clinical-trial or real-world data sets. A list of features and a complete description of the data sets used is provided in Table 1.

### Performance compared with state-of-the-art survival prediction models

To compare the performance of the clinical transformer (including all the aforementioned learning strategies) with those of methods commonly used in the field—specifically, Cox PH, TMB (using TMB as the risk score), and random survival forest, we used five independent data sets (Table 2) from the 12 data sets described in the preceding section.

**Table 2.** Overall performance of the clinical transformer compared with Cox PH, random survival forest models, and TMB.

| **Modeling framework** | **Learning strategy** | **MYSTIC trial (NSCLC)** | **OAK trial (NSCLC)** | **Samstein et al.<sup>25</sup> pan-cancer** | **Thorsson et al.<sup>24</sup> pan-cancer (TCGA)** | **Chowell et al.<sup>3</sup> pan-cancer** | **Chowell et al.<sup>3</sup> evaluated on MYSTIC** |
| --- | --- | --- | --- | --- | --- | --- | --- |
| Clinical transformer | Neural network | **0.670 ± 0.07<sup>a</sup>** | **0.669 ± 0.04** | **0.649 ± 0.02<sup>b</sup>** | **0.734 ± 0.01** | **0.720 ± 0.01** | **0.643 ± 0.004<sup>a</sup>** |
| Linear modeling | Cox PH regression | 0.599 ± 0.03 | 0.620 ± 0.03 | 0.594 ± 0.03 | 0.690 ± 0.01 | 0.709 ± 0.01 | ** |
| Nonlinear modeling | Random survival forest | 0.606 ± 0.04 | 0.664 ± 0.05 | 0.638 ± 0.01 | 0.722 ± 0.01 | 0.714 ± 0.01 | ** |
| Biomarkers | TMB | 0.589 ± 0.05 | 0.543 ± 0.05 | 0.538 ± 0.02 | 0.624 ± 0.01 | 0.550 ± 0.02 | ** |

\*\*Not evaluated.
<sup>a</sup> Chowell et al. pretraining.
<sup>b</sup> GENIE pretraining.

We evaluated the performance of the clinical transformer in a well-defined set of clinical and molecular biomarkers from a study by Chowell et al.,^3^ using concordance index (C-index).^22^ This data set comprises multiple variables that have been integrated into a machine learning model to predict patient response to immunotherapy.^3^ To train our models, we used the same training and testing data splits as in that study, as well as the reported prediction scores from the trained random forest model, along with TMB for comparison against our clinical transformer model (Supplementary Information section A1). In the pan-cancer setting, our clinical transformer achieved a C-index of 0.73, outperforming the Chowell et al.^3^ random forest model, which had a C-index of 0.68, and TMB (which was recently FDA approved for this purpose^23^), which had a C-index of 0.55 (Fig. 2a–c).

**Fig. 2.**
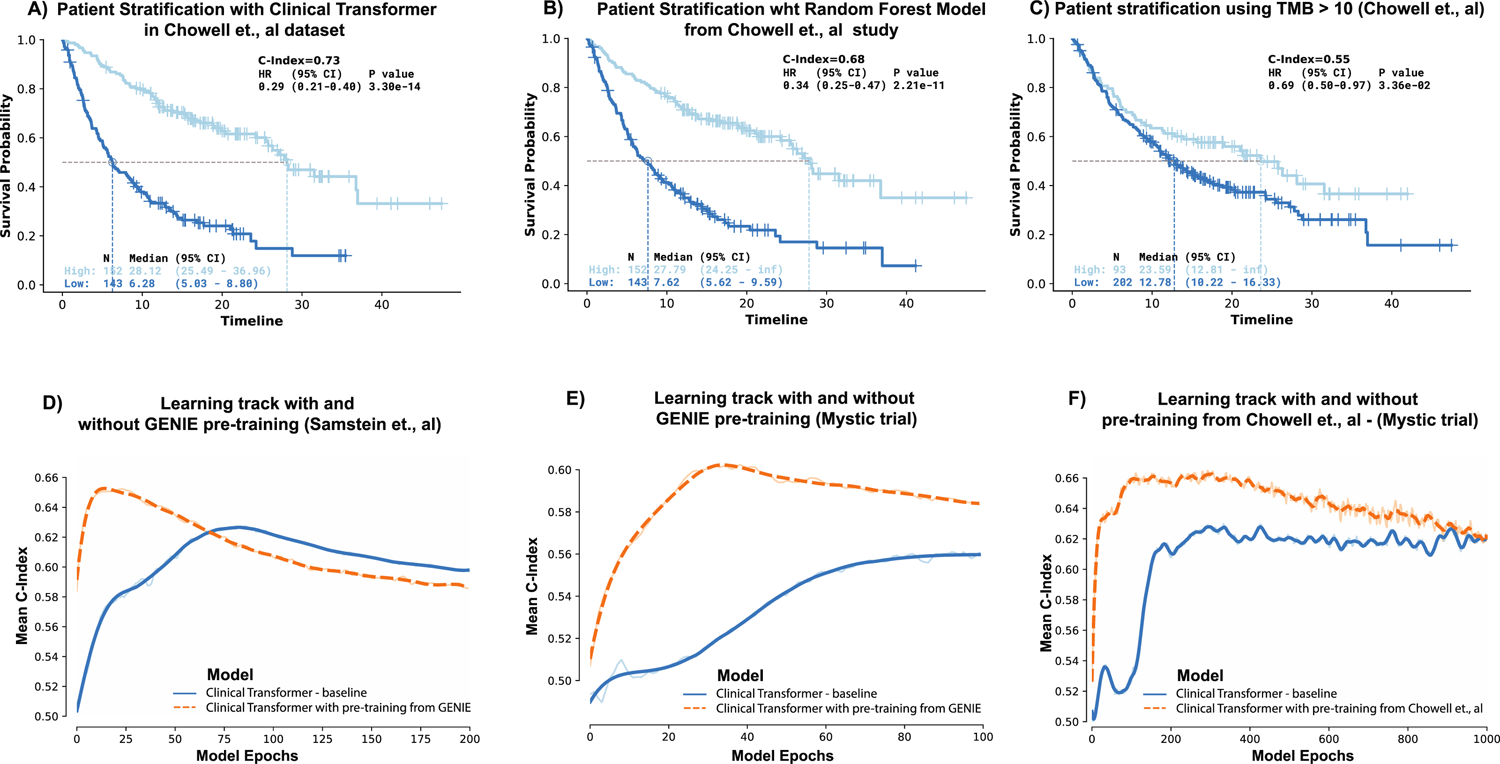
Clinical transformer performance and impact of pretraining. **a**, Performance of the clinical transformer using the median survival score to stratify patients into high and low populations with Kaplan-Meier plots. **b**, Kaplan-Meier curve of Chowell et al.^3^ random forest model used in the testing data set with an optimal cutoff (pan-cancer 0.238). **c**, Kaplan-Meier curves for evaluating TMB score in the Chowell et al.^3^ data set, using 10 mutations per megabase as the cutoff. **d**, Learning curves of the clinical transformer for the training epochs with respect to the C-index for a pretrained model using the GENIE data set and evaluated in the Samstein et al.^25^ pan-cancer data set for the 10 testing splits. **e**, Learning curves of the clinical transformer, comparing the pretrained model using GENIE against the baseline model without pretraining on the MYSTIC data set with 10 testing splits. **f**, Learning curves of the clinical transformer pretrained by using the Chowell et al.^3^ data set and evaluated on the MYSTIC trial with 10 testing splits.

To further evaluate our model’s performance for patient stratification, we assigned patients to either high- or low-risk populations using specific cutoffs. For the Chowell et al.^3^ classifier, we used the optimal cutoff of 0.239 as reported in that study;^3^ for TMB, we used the FDA-approved cutoff of 10 mutations per megabase;^23^ and for our model, we used median score cutoff derived from the training set. Our clinical transformer showed the best patient stratification (Fig. 2a), with a hazard ratio (HR) of 0.29 (95% confidence interval [CI], 0.21–0.40; *P* = 3e-14), compared with the Chowell et al.^3^ random forest model (Fig. 2b), with an HR of 0.34 (95% CI, 0.25–0.47; *P* = 2e-11), and TMB (Fig. 2c), with an HR of 0.69 (95% CI, 0.50–0.97; *P* = 3e*-*2).

To examine the benefit of the gradual learning strategy described in the preceding section, we further pretrained the clinical transformer in a self-supervision mode for 30,000 iterations on the complete Chowell et al.^3^ data set and then fine-tuned it on the survival end point. We then evaluated its performance on an independent data set of 150 patients with non–small-cell lung cancer (74 patients treated with anti-PD-L1 and 76 treated with anti-PD-L1 + anti-CTLA-4 from the MYSTIC trial [NCT02453282]; see Supplementary Information section A2). The clinical transformer showed superior performance when trained on the Chowell et al. data with gradual learning and evaluated on MYSTIC data, with a C-index of 0.643 and an HR of 0.50 (95% CI, 0.34–0.74; *P* = 2.64e-3), whereas the clinical transformer in direct learning strategy showed a C-index of 0.616 with an HR of 0.56 (95% CI, 0.38–0.81; *P* = 3.79e-3). TMB achieved a C-index of 0.608, with an HR of 0.68 (95% CI, 0.47–0.99; *P* = 4.4e-2) (Supplementary Fig. SF2.1, Supplementary Table ST2.1). These results indicate the benefit of the gradual learning strategy in predicting survival outcomes when large data sets are not available.

As demonstrated in Table 2, the clinical transformer outperformed all other approaches. In the independent evaluation of the clinical transformer on IO treatment arms in the MYSTIC study (anti-PD-L1, anti–PD-L1 + anti–CTLA-4; NCT02453282) and OAK trials (anti–PD-1/PD-L1; NCT02008227), we observed survival predictions with C-indexes of 0.67 for MYSTIC and 0.669 for OAK, compared with the random survival forest model, which had C-indexes of 0.606 for MYSTIC and 0.664 for OAK. Cox PH modeling resulted in a much lower performance, with a C-index of 0.599 for MYSTIC and 0.620 for OAK. Similarly, TMB resulted in a C-index of 0.589 for MYSTIC.

Next we sought to examine the ability of the clinical transformer to predict survival based on nonclinical features. We used a set of features reported by Thorsson et al.^24^ to characterize the tumor microenvironment (TME) compiled from TCGA. Thorsson et al.^24^ integrated major immunogenomics methods for the assessment of total lymphocytic infiltrate (from genomic and hematoxylin-eosin image data), immune cell fractions from deconvolution analysis of mRNA sequencing data, immune gene expression signatures, neoantigen prediction, TCR and BCR repertoire inference, viral RNA expression, and somatic DNA alterations. The clinical transformer achieved a C-index of 0.734 and an improved prediction of overall survival (OS) compared with the random survival forest model, which had a C-index of 0.722; the Cox PH model, with a C-index of 0.690; and TMB, with a C-index of 0.624. Finally, we further evaluated the clinical transformer using data from Samstein et al.,^25^ a molecularly derived study in which 1,662 tumors from 32 different cancer types were profiled using the Memorial Sloan Kettering (MSK) Integrated Mutation Profiling of Actionable Cancer Targets (IMPACT) panel (Supplementary Information section A3). The clinical transformer achieved a C-index of 0.649, the random survival forest a C-index of 0.638, the Cox PH a C-index of 0.594, and the TMB a C-index of 0.543. Overall, these results demonstrate the superiority of the clinical transformer in predicting patient survival in a multitude of feature modalities and types, suggesting its potential use in a wide range of clinical bioinformatics applications.

### Use of deep learning by GENIE to predict patient response in small immunotherapy cohorts

“Real-world data” is a broad term applied to data generated in routine clinical practice and has been used to describe a variety of data sets, including UK Biobank,{UK Biobank, #55} Flatiron,^26^ TEMPUS,^27^ and GENIE.^21^ GENIE is a cancer registry assembled through data sharing among 19 leading international cancer centers. These data link clinical and genomic data that can provide useful insights to support the identification of biomarkers associated with patient response to a given treatment. Here, we introduce as part of the clinical transformer framework the use of transfer learning, which is designed to take advantage of the data available in GENIE. As in large language models (e.g., chatGPT), which are pretrained to understand language by using large amounts of data for specific tasks, the clinical transformer uses unlabeled data from real-world evidence to learn general patterns and relationships in the data. The general model is then fine-tuned (or specialized) for a specific task, such as response to treatment or survival, based on a small data set from a clinical trial. We implemented the following three stages to demonstrate the advantage of this approach: (1) pretraining, in which a model is trained by using the GENIE v.11 data set (*N* = 134,626), including 2,290 variables via standard masked self-supervised learning; (2) transfer learning, in which the GENIE model’s weights are transferred to a survival model with the same architecture but outputs a survival score; and (3) prediction of patient survival by fine-tuning the model based on a small cohort of patients with a clear treatment line and survival endpoint. This third stage is benchmarked independently across four IO data sets: Samstein et al. pan-cancer (*N* = 1610),^25^ lung cancer from MSK Multi-modal Integration of Data (MIND) (*N* = 246), the MYSTIC trial (*N* = 325), and the Dana Farber Cancer Institute melanoma data set (*N* = 110) (Table 1). We evaluated direct and transfer learning models by using the C-index and the number of iterations to obtain the peak performance over 10 training (80%) and testing (20%) splits (see Supplementary Information section A3).

In general, transfer learning based on GENIE demonstrated improvement in predicting patient survival across all data sets (average C-index, 0.617) compared with direct learning (average C-index, 0.583) (Supplementary Table ST3.1), with an average reduction in training time of 40% (Fig. 2d, 2e; Supplementary Fig. SF3.1). We further evaluated the advantage of transfer learning on another independent data set taken from the Chowell et al. study^3^ and fine tuned it on MYSTIC trial data (Fig. 2f; Supplementary Information section A4) and obtained a C-index of 0.670, as compared with direct learning, with a C-index of 0.628 (Mann-Whitney-Wilcoxon test, two-sided; *P* = 0.045). For patient stratification, transfer learning achieved an average median risk cutoff, with an HR of 0.49 (95% CI, 0.21–1.15) across 10 testing splits, as compared with the model without transfer learning, which achieved an HR of 0.57 (95% CI, 0.24–1.33).

### Using the clinical transformer’s explainability module to identify features associated with survival outcomes

Clinical applications require information not only about results, such as risk or response prediction, but also about the characteristics and features on which these results are based and their respective underlying assumptions.^28^ Explainability has been an important area of research in recent years^29–31^ to turn black-box models to white-box in order to provide a verification check, increase human confidence in predictions, and unravel the underlying mechanisms behind successful predictions or erroneous results. Here we describe the clinical transformer’s explainability module, which allows for a comprehensive understanding of the factors influencing disease biology and their relationships with patient outcomes.

To estimate feature importance in the model, we used a feature permutation importance algorithm in which the values of a given variable are perturbed and then fed into the model.^32^ The difference between the output C-index from the unperturbed and perturbed feature is then computed. If a feature has a strong effect on the model’s outcome, it will be expected to generate a strong change in the model’s output. Conversely, for features that do not influence the outcome, the change would be close to zero. We evaluated feature importance by 10 permutation tests for each trained model over the testing splits.

In the Chowell et al.^3^ data set, we observed that the most informative features were albumin, neutrophil-to-lymphocytes ratio (NLR), prior chemotherapy, TMB, fraction of copy number alterations (FCNA), and hemoglobin (HGB), whereas human leukocyte antigen (HLA) evolutionary divergence (HED), age, sex, and cancer type did not have a strong effect on the model’s outcome (Fig. 3a). In general, we observed strong agreement between the ranking of the clinical transformer’s feature importance and those in the Chowell et al.^3^ study. To identify the association of the input features with patient response to immunotherapy, we stratified patients into four categories (Fig. 3b), defined by quartile cutoffs based on the model’s predicted survival scores. These resulted in the following patient groups: (1) short-term survivors, patients below the 25th percentile, with a median OS of 4.4 months; (2) mid-low survivors, patients between the 25th and 50th percentiles, with a median OS of 10.3 months; (3) mid-high survivors, patients between the 50th and 75th percentiles, with a median OS of 19 months; and (4) long-term survivors, patients above the 75th percentile, with a median OS of 37 months. Albumin scores demonstrated a statistically significant difference (*P* < 1e-16) between short-term survivors (μ = 3.2 g/L, δ = 0.4 g/L) and long-term survivors (μ = 4.1 g/L, δ = 0.27 g/L) as well as NLR scores, where short-term survivals were observed to have on average high NLR levels (μ = 12, δ = 11.8) and the long-term survivors relatively low average values (μ = 3.15, δ = 1.79) (Fig. 3c). The high variance in the short-term survivors suggests that this group may be less stable than the long-term survivor population. Interestingly, the TMB score exhibited statistical significance only when the long-term survivors were compared with all other populations (*P* < 1e-4). However, there was no statistically significant difference when short-term survivors were compared with intermediate-term survivors (*P* = 0.2) or long-term survivors (*P* = 1.0). Therefore, in accordance with the literature,^25, 33^ high TMB scores (e.g., ≥10 mutations per megabase) could be interpreted as indicative of a positive IO treatment response. Conversely, lower TMB scores may not hold much relevance in predicting the IO treatment response. Moreover, in this data set, the clinical transformer identified approximately 60% of long-term survivors who did not receive chemotherapy prior to IO treatment (*P* < 1e-4; Fig. 3c). Nevertheless, due to the expected overall better outcomes of patients treated with first-line therapies, the association of this factor specifically with IO response is uncertain.

**Fig. 3.**
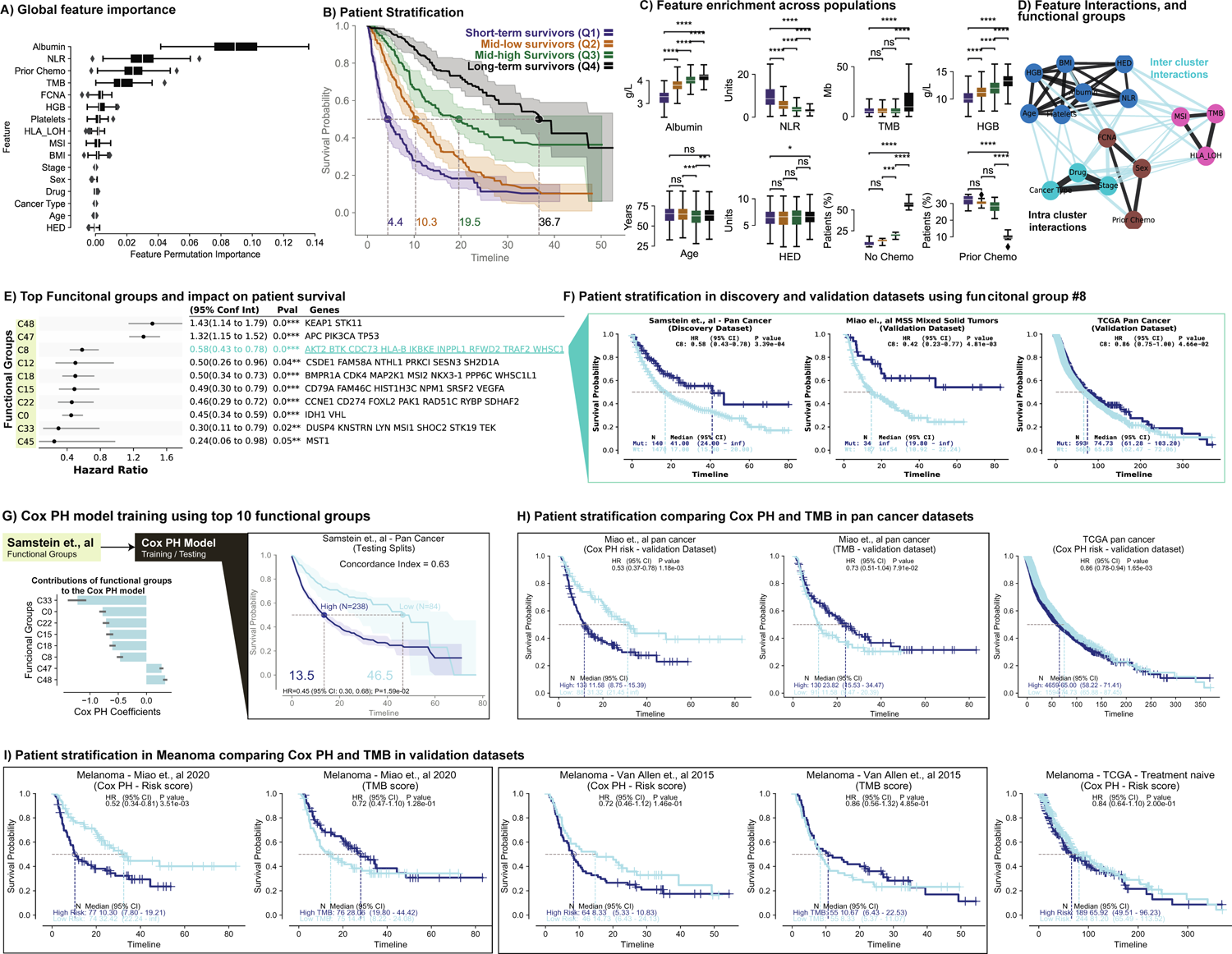
Patient stratification and model interpretability in the test sets. **a**, Global feature contributions across the clinical transformer models using the feature permutation importance algorithm in the Chowell et al.^3^ data set. **b**, Kaplan-Meier curve consistency for different population groups defined by the four quantile cutoffs from the clinical transformer survival scores. Solid lines represent the median survival time and probability from the 10 models on the test splits. **c**, Raw feature value enrichment in the four populations used to stratify the patients. For numerical variables, the y-axis defines the units of each variable (e.g., albumin g/L), whereas for binary features such as prior chemotherapy and cancer type, the y-axis represents the number of patients in each population. Statistical significance from *t* tests is depicted on top of each box plot (*P* value annotation level: ns, 5.00e-02 < *P* ≤ 1.00e+00; *, 1.00e-02 < *P* ≤ 5.00e-02; **, 1.00e-03 < *P* ≤ 1.00e-02; ***, 1.00e-04 < *P* ≤ 1.00e-03; ****, *P* ≤ 1.00e-04). **d**, Feature interaction graph derived from cosine interaction scores. Each color depicts one of the four functional groups identified when clustering the feature pairwise cosine similarities in the test sets. Intracluster (within-cluster) interactions are depicted in black, whereas intercluster interactions (across functional groups) are shown in magenta. **e**, Forest plot depicting the patient stratification for the top 10 functional groups in the Samstein et al.^25^ data set. The majority of functional groups are associated with response to treatment, whereas two groups are associated with poor outcomes. The genes belonging to each functional group are capped at 10 genes for visualization purposes. **f**, Pathway analysis of the functional groups reveal that functional group C8 is associated with innate immune system. Kaplan-Meier curves depict the prognostic value of the group C8 on the discovery data set (training/testing clinical transformer) as well as for an independent data set (Miao et al.^49^) and TCGA data set for patients treated with standard of care. **g**, Cox PH model trained on top 10 functional groups. The bar plot depicts Cox PH coefficients, and the Kaplan-Meier curve shows the performance of the Cox PH model on training/testing strategy. **h**, Evaluation of the Cox PH model in the pan-cancer setting on the discovery data set (Samstein et al.^25^), the pan-cancer IO validation data set (Miao et al.^49^), and TCGA data from treatment-naive patients. **i**, Patient stratification using the Cox PH model derived from the top 10 functional groups on melanoma across independent validation data sets.

### Identification of key functional groups associated with survival outcomes

The attention mechanism is an important component that distinguishes the clinical transformer from other deep neural-network architectures (e.g., fully connected and convolutional neural networks) in generating biological insights. This mechanism enables the model to identify second-order or greater patterns in the data, such as interactions between two features that are associated with patient survival and response. The attention mechanism has been previously employed to interpret model predictions. For instance, in Xie et al.,^34^ Vashishth et al.,^35^ and Vig and Belinkov,^36^ attention weights were used as maps to investigate salient interactions among features. A limitation of these studies was that they considered only the attention network’s weights, which may not provide a direct interpretation and can be influenced by nonlinear relationships within other transformer components (e.g., feed-forward network following multi-head attention, layer normalization). Because these weights may not necessarily correlate with importance values derived from gradient-based methods,^37–39^ they do not fully capture the underlying significance of the features.

We propose a direct approach for assessing interpretability by calculating the similarity between the model’s embeddings, which are located downstream of all the transformer’s neural-network components. We used cosine distance as a similarity measurement between any two feature embeddings (e.g., the embeddings of the independent variables NLR or albumin) as well as the outcome embeddings (e.g., survival) (Fig. 1b). These embeddings encode both linear and nonlinear relationships between input features in the context of clinical outcomes. The cosine similarity between two features provides insights into their relationship. A high cosine similarity suggests that the features share similar information and may be redundant. Conversely, a cosine similarity close to zero indicates that the features are independent and encode different information, such as orthogonality. A negative cosine similarity signifies that the two embeddings encode inversely. With this approach, we can make use of the information flow through the entire transformer network as opposed to independent attention scores derived from individual attention heads upstream in the neural-network model.

Applying this approach to the Chowell et al.^3^ data set showed albumin and TMB to be independent features with the highest level of orthogonality (cosine similarity of 0.09; Supplementary Table ST2.1, Supplementary Fig. SF6.1). This result suggests that albumin and TMB contribute complementary information to the prediction of patient survival, consistent with previous studies showing that albumin has a positive effect on predicting patient response to immunotherapy when combined with TMB.^40^

Input features were grouped into four functional groups based on their cosine similarity (Methods), representing groups of features that shared similar information with respect to the survival endpoint (Fig. 3d). Cluster 1 included a grouping of TMB, MSI score, and HLA–loss of heterozygosity (LOH), with a mean cosine similarity of 0.65 (0.59, 0.68, and 0.70 cosine similarities for MSI-TMB, HLA-LOH–MSI, and HLA-LOH–TMB interactions, respectively) (Supplementary Table ST6.1), implying that these three features may share a common underlying mechanism. It has been well documented that a high MSI score is associated with high TMB,^41–44^ which provides a valid rationale for the grouping of these features. One possible link between these two features and HLA-LOH is the process of antigen availability and presentation. TMB and MSI are both surrogates for neoantigen availability, and tumors with higher neoantigen loads are under greater pressure to lose the ability to present those neoantigens through the loss of HLA. Cluster 2 consisted of patient-related variables, including albumin, body mass index, HED, HGB, NLR, platelets, and age. Interestingly, although HED is a molecular-based biomarker, it was clustered together with clinical laboratory patient-level test markers, rather than with TMB, MSI, or HLA-LOH, which are also molecular markers but are directly derived from the tumor. This distinction can be attributed to the fact that HED is derived from germline DNA and represents an immune characteristic at the patient level. Cluster 3 included FCNA, prior chemotherapy, and sex, with a mean cosine similarity of 0.60. Finally, cluster 4 had the highest average within-cluster cosine similarity, suggesting that cancer type, drug class, and tumor stage included in this cluster shared similar information (cosine, >0.95) with respect to the clinical endpoint. Nevertheless, this finding may reflect the underlying dependencies among drug, cancer type, and stage of disease that are associated with clinical practice.

To further explore an application of the explainability module, we investigated the tumor biology and its association with survival based on mutational profiling in the Samstein et al. data set.^25^ Here, we formed clusters on the basis of cosine similarity, the tumor tissue gene-level mutational features, to identify only molecularly derived functional groups associated with patient survival with immunotherapy (using the MSK IMPACT panel of 469 genes) (Supplementary section A5). Mutational features were clustered into 50 functional groups and were ranked on the basis of their association with benefit of immunotherapy (Fig. 3e, Supplementary Fig. SF5.1). We aggregated each functional group of multiple genes (each gene is a binary variable with an indication of 1 to denote that it is mutated and 0 if not) to a single variable using a simple rule, as follows: if any gene in a given functional group is mutated, the representing variable of that functional group is assigned the value 1; if none of those genes is mutated, it is assigned 0. Two functional groups, denoted C47 (HR = 1.32; *APC*, *PIK3CA*, and *TP53*) and C48 (HR = 1.43; *KEAP1* and *STK11*), were associated with short-term survival with immunotherapy. Indeed, multiple studies have shown the negative prognostic impact of *KEAP1* and *STK11* for both immunotherapy and chemotherapy.^45–48^ Alternatively, we evaluated the survival impact of the functional group C8, which was composed of the genes *AKT2*, *BTK*, *CDC73*, *HLA-B*, *IKBKE*, *INPPL1*, *RFWD2*, *TRAF2*, and *WHSC1* and is associated with regulation of the immune response (GO:0002682, *P* = 8.69e-4). We used this group to stratify patients across independent IO-treated data sets, resulting in a significant and selective benefit of IO treatment. We observed a meaningful stratification for both Samstein et al.^25^ and Miao et al.^49^ pan-cancer IO-treated patients in these data sets (HR = 0.58, *P* = 3.3e-4; HR = 0.42, *P* = 4.8e-2, respectively), whereas no meaningful stratification was observed for pan-cancer non–IO-treated patients in the TCGA data set (HR = 0.86, *P* = 4.6e-2; Fig. 3f).

### Translating the clinical transformer to simple and interpretable linear models

As described in the preceding section, the clinical transformer was able to extract functional groups by clustering the features on the basis of the cosine similarity of their outcome embeddings. We sought to examine whether these patterns could be leveraged to construct a simple linear model. To this end, we binarized each functional group by using the strategy just described, in which each functional group is represented as a binary variable describing the presence of a mutation in any of the given genes. This single representing variable was then used as input to a Cox PH model to predict patient OS. Multiple function groups could be also used by representing each one as a single variable to construct a multivariate Cox PH model. Therefore, we selected the top 10 molecularly derived functional groups identified in the Samstein et al.^25^ data set (those showing the strongest relationship with OS; Fig. 3e) as inputs to a multivariate Cox PH model to predict patient OS (see Methods). Intriguingly, we observed only a small decrease in performance with this approach compared with the complex clinical transformer model. The Cox PH model resulted in an average C-index of 0.63 (HR = 0.53; Miao et al. data set^49^) (Fig. 3h), whereas the clinical transformer with the whole set of features (including clinical and demographics) resulted in a C-index of 0.65. In contrast to Cox PH and random Cox PH, the clinical transformer guided the selection of the functional groups, and the performance could not be recovered by using a random selection of functional groups (C-index = 0.55 on test data set, HR = 0.76; Miao et al. data set^49^) or a random set of 10 hallmark gene sets (C-index = 0.58 on test data set, HR = 0.76; Miao et al.^49^ data set) (Supplementary Fig. SF5.2). Stratifying the population by the median risk score cutoff derived from the discovery data set (Samstein et al.^25^), the Cox PH model achieved an average HR of 0.45 across the testing splits (Fig. 3g). We evaluated the model against several independent validation data sets and obtained an HR of 0.53 in the pan-cancer IO-treated population from the Miao et al.^49^ data set, which outperformed TMB (using median cutoff) (Fig. 3h), as well as in the non–IO-treated patients in TCGA data set, where the Cox PH model did not show any significance for stratifying patients (HR = 0.99). We furthered evaluated the functional-group Cox PH model in melanoma, where we observed the same trend across all immunotherapy validation data sets and the opposite trend for TCGA melanoma (Fig. 3i). Note that our functional-group Cox PH model achieved an HR of 0.52, outperforming TMB with an HR of 0.72 in the Miao et al. data set.^49^ The same pattern was observed in the Van Allen et al.^50^ data set, where the Cox PH model achieved an HR of 0.72, outperforming TMB with an HR of 0.86 (Fig. 4i).

**Fig. 4.**
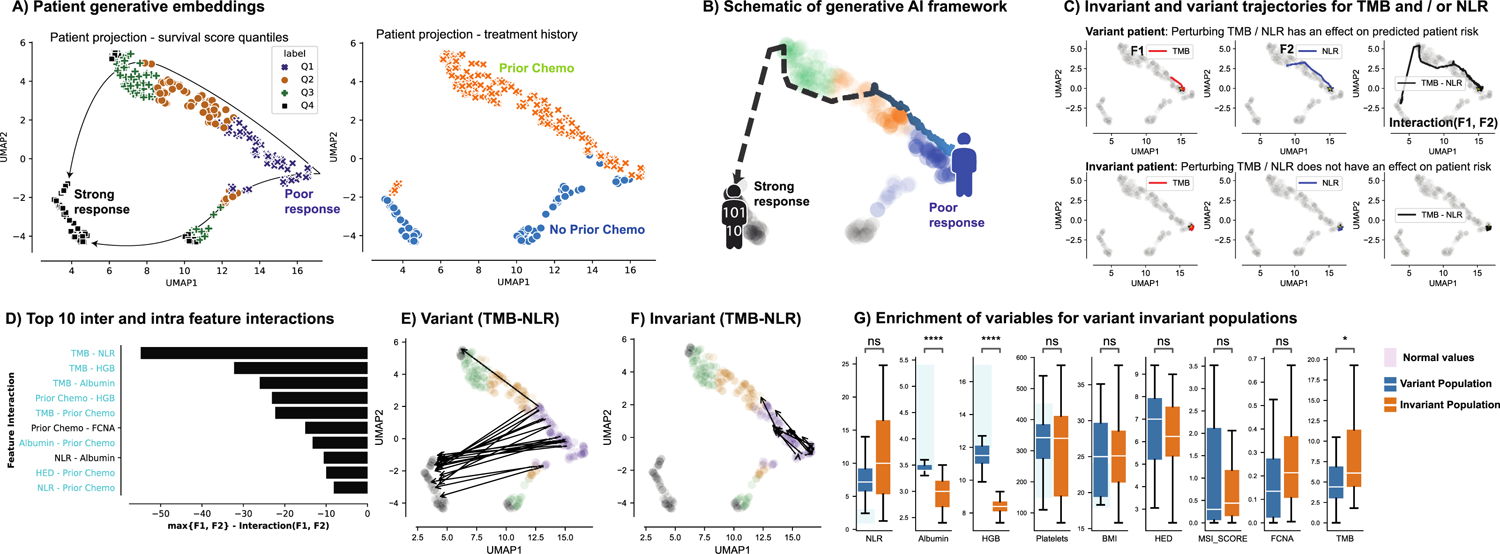
Impact of functional groups on patient response to IO treatment. **a**, Two-dimensional projection of patient embeddings with UMAP labeled by response of patient populations (left) and by prior chemotherapy (right). Each point in the UMAP plane is a patient’s embeddings projection. **b**, Schematic of perturbation framework and expected survival trajectory in the latent space of patients under perturbation. **c**, Response trajectory for the patients who, under perturbation of the top feature interaction (TMB-NLR), moved from poor-response to super-response (variant population). **d**, Top inter- and intra-feature interactions using the generative module and measuring the impact of perturbing one and two features at a time and calculating the maximum effect of the perturbation for individual and paired features. **e**, Example of the variant population (patients showing a change in survival score) under TMB and NLR perturbations. **f**, Example of an invariant population, in which a patient survival score is independent of the perturbed features. In this case, for TMB and NLR perturbations, these patients exhibited no change in survival score. **g**, Differences in clinical and molecular features for the variant and invariant populations.

The results presented in this section demonstrate a valuable opportunity to translate patterns recognized with complex nonlinear models into an easily explainable linear model such as the Cox PH. This approach has the potential to facilitate a deeper understanding of the relationships between input features and outcomes and accelerate the discovery of a composite biomarker to be translated to companion diagnostics.

### Clinical transformer embeddings capture of biological patterns associated with response

As in language models, which generate word embeddings that can incorporate both word-level characteristics and contextual semantics,^11, 51–53^ the clinical transformer’s outcome embeddings may incorporate relationships among the molecular and clinical features of a patient. These relationships are encoded in the context of the clinical endpoint (e.g., survival time) that the model was trained to predict and could potentially reflect higher-order dependencies required to unravel biological mechanisms related to response, resistance, or survival. Therefore, we transformed the 128-dimension outcome embeddings space (Fig. 1b) to a two-dimensional space by using the Uniform Manifold Approximation and Projection (UMAP) transformation, a method that is widely used in many computational biology applications.^54, 55^ Fig. 4a presents an example of this projection for the patients in the Chowell et al.^3^ data set. Clustering and labeling of outcome embeddings were projected into this UMAP plane, enabling us to explore patterns in the data in the context of response level, treatment lines, and other features of interest.

As an example, Fig. 4a demonstrates how patients with specific clinical features, such as survival and treatment history, are organized in a clinically meaningful order on the embeddings plane of the clinical transformer. The left panel in Fig. 4a shows a clear trajectory on the embeddings space from short-term survivors to long-term survivors for IO-treated patients in the Chowell et al.^3^ data set. The right panel in Fig. 4a demonstrates a clear separation of two distinct patient groups, one with prior chemotherapy and the other without.

### Exploring alternatives of clinical response for patients with perturbation analysis

Next, to examine potential alternatives to response of short-time survivors, we selected patients with a similar clinical response (e.g., short-time survivors) and separately perturbed each of their input features (i.e., artificially changing the value of a feature such as NLR, TMB, or gene or pathway expression) while maintaining the other features fixed. By calculating the difference between the predicted survival score of the patients before and after perturbation (see Methods and Supplementary section A7 for more details), we were then able to identify which patients within the same original clinical response segment may potentially have improved survival based on the expected pathways that were modulated (the perturbed pathway is related to the mechanism of action of the therapy investigated). Through examination of the other features of these patients, we may be able to identify new segments for potential treatment, and by examining patients whose disease failed to improve, we may inform potential resistance mechanisms and combination opportunities.

Individual feature perturbation (Fig. 4c) showed that patients can be sensitive to one, two, or none of the perturbed features, highlighting the sensitivity of a given patient to the input features. Fig.4c (top) shows an example of a patient who was more sensitive to changes when TMB and NLR were perturbed together rather than individually. Fig. 4c (bottom) depicts a patient who was not sensitive to NLR or TMB perturbations, individually or together. Next, we exhaustively perturbed all pairwise combinations of features to observe whether any two features resulted in an impactful interaction on the perturbed patient’s outcome. Interestingly, we found that interactions between features from different clusters (i.e., clusters indicated in Fig. 3d) had a stronger impact on a patient’s survival than interactions between features from the same cluster (binomial test between clusters, *P* = 0.004; within clusters, *P* = 0.58; Fig. 4d). As demonstrated in Fig. 4d, TMB-NLR exhibited the most impactful pairwise-feature interaction, followed by TMB-HGB and TMB-albumin. Importantly, we identified two population subtypes, one that exhibited a strong change in survival score when either of those features were perturbed (hereafter denoted as the variant population; Fig. 4e) and another that was invariant to perturbations (hereafter denoted as the invariant population; Fig. 4f). We further investigated the patients that exhibited improvement of their predicted survival from short- to long-term survival (patients labeled Q1 by using the cutoff defined from the training set). As expected, patients from the variant population exhibited values within the normal ranges for albumin, HGB, and platelets, as indicated in Fig. 4g (blue boxes) (albumin, 3.4–5.4; NLR, 1–3; HGB, 12–17; platelets, 150–450) (Fig. 4g). These values may serve as indicators of better health compared with patients with values outside of the normal ranges. As expected, high values of TMB along with low values of NLR tended to produce the greatest changes in survival scores for this population (Fig. 4g). In contrast, the invariant population tended to be in worse health, with albumin and HGB outside the normal range (Fig. 4g).

### Identification of potential drivers of response and resistance to immune checkpoint inhibitor treatment via perturbation of a T-cell gene expression signature

The clinical transformer was used to train a model of survival for patients with melanoma (skin cutaneous melanoma [SKCM], IO treatment naive) in TCGA based on the gene signatures of the TME defined in Bagaev et al.^56^ Quartile survival groups resulting from this training are illustrated in Fig. 5a.

**Fig. 5.**
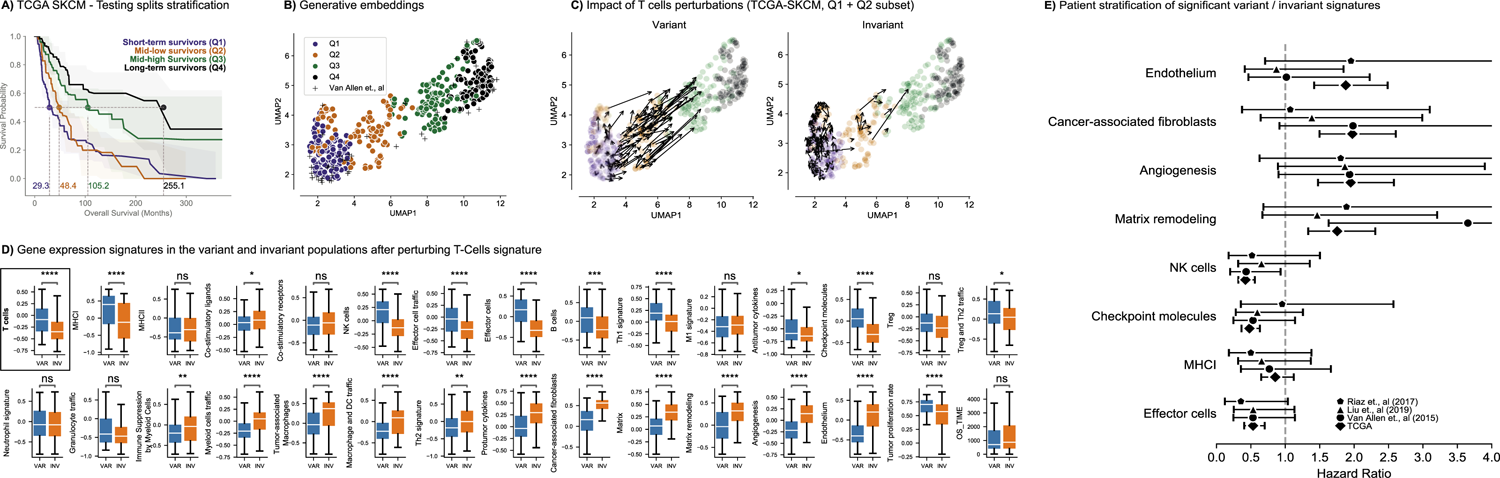
Clinical transformer and TME on SKCM data. **a**, Survival stratification of SKCM patients from TCGA. Patients are grouped into quartiles from the clinical transformer survival score. The solid line represents the mean survival time across the 10 testing splits. **b**, UMAP projection of patient embeddings in the TCGA SKCM data set. **c**, Effect of T-cell perturbations (variant and invariant populations) in TCGA SKCM data set exclusive to patients in the Q1 and Q2 populations. The size and direction of the arrows reflect the effect and directionality, respectively, of the perturbation. **d**, Distribution of TME signatures for the variant and invariant populations as an effect of T-cell perturbations. **e**, Forest plot of top gene signatures associated with response and resistance in TCGA and IO-treated populations.

Having established a model of the TME’s impact on survival in melanoma patients (see Supplementary Figs. SF8.1, SF8.2 and Bagaev et al.^56^), we sought to determine whether such a model could be used to better understand or predict drivers of response and resistance to checkpoint blockade in this setting. Fig. 5b presents the UMAP projection of the generative embeddings of the patients from this model. We then perturbed the values of a T-cell gene expression signature within the Q1 and Q2 patients of the TCGA-SKCM cohort to mimic the increase in T-cell infiltration and activation that would be expected upon checkpoint blockade. As with the example in the preceding section, this perturbation generated a variant group of patients, for whom an increase in T-cell gene expression mediated improved survival, and an invariant group, for whom it did not. The changes of these perturbations for the variant and invariant groups are shown on the generative embeddings UMAP plane in Fig. 5c. A comparison of gene expression signature differences between these variant and invariant groups is presented in Fig. 5d (Supplementary Fig. SF8.3). The variant group of patients, who might be considered to potentially benefit from checkpoint blockade, demonstrated significantly increased levels of expression for signatures associated with major histocompatibility complex class I (MHC-I) and effector immune cells, suggesting, as has been described elsewhere,^56^ that a degree of preexisting immunity to the tumor, together with functional antigen presentation, are key determinants of benefit from checkpoint blockade. In contrast, the invariant population of patients, who might be considered to be resistant to checkpoint blockade, demonstrated increased signatures of angiogenesis, matrix remodeling, and infiltration of both cancer-associated macrophages and fibroblasts. Suppression of T-cell immunity by intratumoral macrophages through a number of mechanisms has been well described as a mechanism of resistance to checkpoint blockade, as has the exclusion of T cells from the microenvironment by both cancer-associated fibroblasts and changes in the extracellular matrix. The impact of angiogenesis on response to checkpoint blockade is less well established but might also be expected, given the necessity of a functional blood vessel system to enable access to the tumor by T cells.

The expected and interpretable nature of the potential mechanisms of response and resistance suggested by the clinical transformer in this context are encouraging with respect to the ability of this approach to yield biological insights. As a final step, we sought to further validate these findings by assessing the ability of the identified signatures to stratify melanoma patients treated with checkpoint blockade. Gene signatures were ordered with respect to the magnitude of difference they demonstrated between variant and invariant populations (Supplemental Table ST8.1). The four signatures with the most significantly increased associations in the variant population were effector cells, MHC-I, checkpoint molecules, and natural killer cells, and the four signatures with the most significantly decreased associations were endothelium, cancer-associated fibroblasts, angiogenesis, and matrix remodeling. These eight signatures were examined in data from three studies.^50, 57, 58^ From these cohorts we selected patients who had a second line of IO treatment and whose tumor biopsies were obtained prior to IO therapy. Data from all three studies were initially pooled to define a global median for each of the signatures. Patients in each study were then stratified for survival around the mean for each signature.

Results are shown in the forest plot in Fig. 5e. Although none of the differences between high and low groups with respect to stratification of survival reached statistical significance, several signatures showed consistent trends for differential HRs across studies. For example, high vs. low levels of the effector cell signature resulted in HRs of 0.53, 0.53, and 0.35 for survival across the three studies, respectively, whereas matrix remodeling produced HRs of 3.65, 1.46, and 1.89 (Supplementary Table ST8.2). These results speak to the potential for the clinical transformer to provide insights with respect to response and resistance, even with early clinical data that do not include the treatment under investigation.

## DISCUSSION

Modified transformer architectures have enabled a significant acceleration in the development of computational models that make meaningful predictions from complex and heterogeneous biological and clinical data sets. The self-attention mechanism in transformer architecture, which enables the capture of dependencies among all input features, makes transformers highly appealing models for survival analysis. This is particularly due to the wide range of molecular and clinical features that play an important role in influencing patient outcomes and treatment response. Survival analysis is an area that has been dominated by statistical approaches, such as multivariate modeling using Cox PH for predicting individual patients’ responses to therapy based on their genomic and clinical features. Several nonlinear approaches have also been proposed, including gradient boosting machines, among others. In recent years, deep neural networks have also been applied to survival analysis. For instance, Katzman et al. developed DeepSurv, a personalized treatment recommender system using a Cox PH deep network^59^. Yousefi et al. implemented SurvivalNet, which uses Cox partial-likelihood to train a neural network to predict survival outcomes.^60^ Hu et al. developed a transformer-based survival model that utilizes ordinal regression to optimize survival probabilities over time.^61^ However, none of these methods consider the interactions among the features in an explicit way as part of the model or enable reviewing of the model at the single-patient level.

We developed the clinical transformer framework, a deep-learning model with explainability that can predict survival outcomes for different lines of treatment with greater accuracy than current state-of-the art prediction models. We demonstrated how the clinical transformer is trained in a self-supervision mode on TCGA or GENIE public data sets (regardless of the clinical endpoint) and is then fine-tuned to make improved predictions on specialized tasks, such as survival prediction for smaller data sets (e.g., clinical trials). This approach allowed us to leverage the wealth of information in large data sets, including unlabeled data, to enhance predictions on limited data sets for which response and survival end points are available. Basing our analysis on several independent data sets, including different input modalities, cancer type, and lines of treatment, we found that the clinical transformer could capture low- and high-order relationships that are encoded within the variation in the clinical, demographic, and molecular data of patients. Importantly, using cosine-distance between the embeddings of the input features, we successfully used the clinical transformer to discover patterns in the data (e.g., interactions between features), such as separated clusters for patient health and immunogenicity. Using the cosine-distance also enabled us to map the dependencies between the input features associated with survival and to recommend minimalistic linear models with a small number of variables, which could be more easily translated to clinical practice and may introduce new insights to response and resistance. For example, our clustering analysis, based on pairwise cosine-distance between the input features, identified two functional groups, C47 (HR = 1.32; *APC*, *PIK3CA*, and *TP53*) and C48 (HR = 1.43; *KEAP1* and *STK11*) associated with poor response to immunotherapy. The *KEAP1* and *STK11* genes are widely known for their role in immunotherapy resistance, illustrating the potential clinical utility of feature-based identification of molecular functional groups. We then showed that these molecular functional groups could be represented as binary variables (i.e., 1 for a mutation, 0 if not) and used as inputs to construct a Cox PH model to analyze the role of molecular features in patient OS. When evaluated against independent validation data sets, we showed that our functional-group Cox PH model outperformed standard methods in pan-cancer IO-treated patient data sets.

Another essential function of the clinical transformer, particularly in early clinical studies with no treatment data available, is perturbation analysis. Using data sets of patients before IO treatment was accessible in the community (e.g., TCGA), we selected patients with short-term survival and perturbed specific molecular features that were expected to change in the TME due to IO treatment. Subsequently, we predicted their survival outcomes. This process allowed us to identify two patient populations: those whose survival improved and those whose survival did not. By further investigating the TME profiles of these patients, we were able to pinpoint factors associated with response and resistance to IO treatment, thereby supporting the potential of this approach to identify patient populations that could benefit from IO treatment based on early data lacking such treatment information.

The current version of the clinical transformer is designed to handle only a few hundred input features effectively (e.g., requiring RNA gene expression data to be aggregated into signatures before training a model). Additionally, for effective performance on relatively small data sets, it is necessary to pretrain the clinical transformer in a self-supervision mode on thousands of patient entries with a similar set of features. Unsurprisingly, the performance and interpretability of the clinical transformer rely on the quality and perspicuity of the features (e.g., gene expression signatures) it uses. We observed that even when prediction performance is high, the lack of direct interpretability in certain features (e.g., immune landscape of cancer signatures^24^) hinders the generation of actionable insights in the relevant clinical context. Conversely, well-defined clinical features^56^ that are structured on the basis of melanoma TME demonstrated highly preferential performance for that indication compared with others. As with other machine learning and AI applications, careful review of the clinical transformer output in the correct context is required to avoid misinterpretation (e.g., prior chemotherapy treatment is probably due to the expected overall better outcomes of first-line patients rather than the association of this factor specifically with response to IO treatment). A key assumption in the perturbation analysis is the presence of sufficient natural variation in patient biology and survival outcomes, allowing the model to learn this variation effectively. This assumption is dependent on the dimensionality and complexity of the data and study, as well as the size of the data set. Additionally, the generative nature of the perturbation analysis in the clinical transformer may result in “hallucinations,” making it essential to validate findings on independent data sets and carefully examine the results.

A key advantage of deep neural networks over conventional machine learning is their flexibility in designing model architecture that can learn from diverse types of data sets. These include both flexibility in including different input features (e.g., molecular signatures and clinical features) and target functions (e.g., prediction of progression-free survival or response). With the proliferation of available biological data from the multitude of assays in preclinical and clinical settings, the digitization of clinical samples, and the vast body of knowledge that is available in the literature, our ability to integrate these data to understand disease biology and response and resistance to treatment is highly valuable. The clinical transformer framework presented in this study is a promising step in this direction. We expect that this research will benefit the scientific community by fostering further developments in maximizing data utilization for the benefit of patients.

## METHODS

### Model architecture

The clinical transformer is composed of three main components: (1) an embedding layer that projects the input features *F* ∈ R*^N^*^×1^ to an embedding space *E* ∈ R*^N^*^×*dk*^ by using the feature name and value pairs; (2) a transformer encoder composed of *l* number of layers that transforms the input embeddings *E* into the output embeddings *P* ∈ R*^N^*^×*dk*^, which encodes feature interactions via the self-dot product attention; and (3) a prediction layer that uses the output embeddings *P* from the transformer encoder to perform the training task (survival or self-supervision) via different loss functions.

### Input embeddings layer

The initial transformer architecture proposed by Vaswani et al.^11^ uses positional encoding vectors to account for the absolute location of tokens in the sequence. Because we did not use sequential data in our work, we excluded the positional encoding vectors from the transformer architecture. The input feature vector **x**, composed of continuous and categorical features, is treated as a set of key value pairs **x** ≡ {**x***_k_*,**x***_v_*}, where each element {*x_i_* ≡ *x*_(*i*,*k*)_,*x*_(*i*,*v*)_} consists of a feature name *k* and its corresponding value *v*. To fit into this encoding schema, categorical variables are converted to ordinal arrays (e.g., *n* categories converted to *n* − 1 integers).

Feature names **x***_k_* ∈ R*^N^*^×1^ are embedded into a latent representation *E_k_* ∈ R*^N^*^×*d*^*^k^* via a standard text embedding layer, while feature values **x***_v_* ∈ R*^N^*^×1^ are projected into a latent space *E_v_* ∈ R*^N^*^×*d*^*^k^* by plugging them into a dense layer. Therefore, each component of the feature value space is decomposed into a linear combination of the learned weights. The aggregated embeddings *E* = *E_k_* + *E_v_* are the input into the transformer layer (Fig. 1b).

Encoding the input feature names and values enables the model to utilize a large and diverse range of features from different modalities, which is common in computational biology. This embedding schema differs from previous approaches that have used transformer models for tabular data, where inputs are fixed-length vectors that are limited to a given number of features.^16–18^ In other words, it is not necessary for our clinical transformer to have all features available as input, which can be useful when working with missing data and reduces the complexity of the model on highly sparse data sets.

### Transformer

The clinical transformer encoder is a multilayer bidirectional transformer (similar to BERT) that is based on the original implementation from Vaswani et al.^11^ The transformer consists of *l* blocks containing a multi-head self-attention network, a position-wise feed-forward layer with element-wise addition with a layer normalization. The core of the transformer is the self-attention layer that enables the model to selectively focus on relevant features from the input space by identifying similarities among the input features while associating those similarities with the model outcome. Formally, input embeddings *E* are projected into three parametric matrices, the Key (*K*), Query (*Q*), and Value (*V*). Queries represent current information for each input feature, and keys represent the information to which features will be attending. The output of the attention mechanism is defined as the Softmax function of the product between *Q* and *K^T^*, normalized by a *d_k_* and multiplied by the *V* matrix as follows:

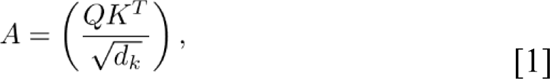

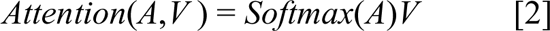

where *Q*,*K*,*V* ∈ R*^N^*^×*d*^*^k^*. *N* is the number of input features and *d_k_* is the dimension of key, query, and value vectors. The attention matrix *A* ∈ R*^N^*^×*N*^ captures contextual similarities between queries and keys. The output embeddings of the transformer encoder *P* ∈ R*^N^*^×*d*^*^k^* are retrieved from the attention matrix after the pointwise and normalization layers.

### Prediction layer

The clinical transformer supports two learning modes: (1) time-to-event prediction (survival analysis) to predict patient survival with a given treatment; and (2) unsupervised learning, in which the model identifies feature interactions by looking only at the input features (Fig. 1b), that is, self-supervision. Similar to BERT, we also included the special features [CLS], [MASK], and [PAD] to represent the objective task (survival), masked input features (for unsupervised learning), and unavailable features (for padding absent features), respectively.

#### Survival task

The final hidden state of the special task feature *P*^[CLS]^ ∈ R^1×*dk*^ is used as the aggregate feature representation of the input data (described as patient embeddings). This vector is passed through a single neuron layer without bias parameter and with weights *W* ∈ R^1×*dk*^. Thus, the survival output score is a scalar defined as β = *P*^[*CLS*]^ ∗ *W*^*T*^ with a linear activation function.

To optimize model parameters toward patient survival outcomes, instead of a binary response or text translation, we used the concordance metric in the survival analysis workflow as a measure of model discrimination. Harrell’s C-index is defined as the proportion of observations that the model can order correctly in terms of survival times.(^62^) The C-index can be interpreted as a generalization of the area under the receiver operating characteristic curve that considers censored data. It represents the global encapsulation of the model’s discrimination power and its ability to provide a reliable ranking of survival times based on individual risk scores. In our workflow, the concordance-based model discrimination was implemented by using a loss function with a sigmoid approximation of Harrell’s C-index. This led to an objective of the form:

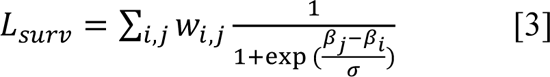

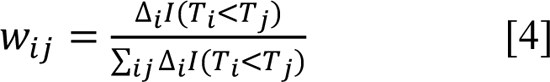

where the indices *i* and *j* refer to pairs of observations in the data, Δ*i* = 0 if censored and 1 if deceased, *T* is the corresponding survival time, β is the predicted survival scores from the clinical transformer, and *σ* is a smoothing parameter for the sigmoid approximation.

#### Masked pretraining task

Pretraining a transformer model has proven to be an effective strategy to leverage relevant patterns directly from the raw data in the absence of labels. In the pretraining mode of the clinical transformer, 20% of the input feature names are randomly replaced by the special tag [MASK] while its original value is unchanged (similar to language model pretraining). The model is then trained to predict the masked feature names and values by using as input the unmasked features and their respective values (Fig. 1b). The input feature vector is composed of masked and unmasked features and is passed through the clinical transformer to obtain the output embeddings *P*^[*MASK*]^ ∈ R^*F*×*dk*^. These vectors are fed to a dense layer with a Softmax activation function to predict the original masked feature names. The output embeddings *P*^[*MASK*]^ are also fed into another dense layer with a linear activation function and outputs the masked feature values. To optimize the model, we used standard categorical cross-entropy for predicting the masked feature names (*LOSS*_*names*_) and the mean square error loss to predict the masked feature value (*LOSS*_*values*_). The final loss corresponds to the weighted sum of the independent losses (α_1_ ∗ *LOSS*_*names*_ + α_2_ ∗ *LOSS*_*values*_), where *α*_1_ = 1 and *α*_2_ = 0.01 are set as default parameters and define the contribution of predicting the name and value. In our current settings, we prioritize the prediction of feature names, given that masked values are also included as inputs. The clinical transformer framework is also designed to support custom loss functions.

#### Feature position invariant trick

To encode feature names and values for tabular data, it is critical for the model to ignore the position where the features are placed. To make the model position invariant, we removed the position encoding layer. In addition, to force the model to avoid any association between position and outcome, we used a simple trick in which the order of the input features is randomized for each input sample during training (except for the [CLS] feature, which is always the last feature). This processing forces the model to ignore the position where features may occur.

### Model training and evaluation

Motivated by the fact that IO-related clinical data with patient outcomes are difficult to obtain in high numbers, we sought a strategy to enable the use of all available clinical data sets to maximize the value of limited IO clinical data. Our framework can leverage the use of other clinical data sets, even with no outcome labels, to improve predictions in smaller clinical data sets. To this end, we evaluated three training strategies. (1) In direct learning, a machine learning model is trained to predict a target task, given an input data set. (2) Gradual learning consists of an intermediate step in which a model is trained over unlabeled data taken from the input data set before training over the target task. In natural-language processing, this is done by predicting a word that is masked in the original text (self-supervision). The obtained model is then transferred to a new model designed to predict the target task of interest. In gradual learning, the same data set used for pretraining is used for fine-tuning (e.g., pretrain on entire data set and fine-tune survival on entire data set). (3) Finally, transfer learning, which is used when a large data set is available, entails pretraining a large model using all unlabeled data and fine-tuning on a target data set and task (e.g., pretrain on the GENIE data set and fine-tune survival prediction over the Samstein et al.^25^ data set).

### Explainability framework

#### Cosine similarity between patient embeddings

We defined “post-attention” as the pairwise cosine similarity among all features in their latent representations. Formally, for each pair of vectors *P*^[*fk*]^,*P*^[*fl*]^ ∈ R^1×*dk*^ from output embeddings, we computed the cosine similarity score as:

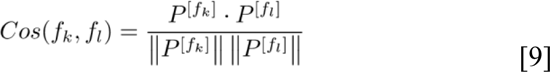

where *f_k_* and *f_l_* are any two features present in the input data point *x_i_*. The complete pairwise feature similarities can be depicted as the square matrix *S*(*x_i_*) ∈ R^*F×F*^. The rationale for using the output embeddings from the last encoder layer is that those embeddings preceded the outcome classifier networks (survival, masked prediction). These embeddings represent the aggregated information of the input feature interactions in the sample *x_i_*.

Given that the outcome embedding vector *P*[*CLS*] ∈ R^1×*dk*^ is directly associated with the outcome variable (because this vector is used as input to the prediction layer), the cosine similarity between this vector and any feature vector *P*^[*fk*]^ reflects the contribution of the feature *fk* to the outcome. Thus, for a given input data point *x_i_*, we can identify and rank the features, encoding the similar information to the predicted outcome by computing all their cosine similarity scores to the outcome embedding.

Similarly, the cosine similarity among feature embeddings *P^Fl^, P^Fk^* ∈ R^1×*dk*^, where *F_l_*, *F_k_* ≠ [CLS], depicts local relationships and feature interactions. Similar to language models, in which two words can describe semantic similarities within a certain context, the cosine similarity between two features describes their information content and can be seen as a local feature interaction.

#### Functional groups

A functional group ζ is defined as the collection of interacting features encoding related information on a global scale. To obtain functional groups, we averaged the pairwise similarities *S*(*x_i_*) ∈ R^*F*×*F*^ across a given population (e.g., an entire population or specific populations) and clustered them, using agglomerative clustering, k-means, or any clustering algorithm that can better represent the data, with a predefined number of ζ*_n_* clusters. For selecting the best number of clusters, the elbow or silhouette score can be applied. To perform clustering analysis, missing pairs are encoded with 1 value.

#### Functional group ranking

In a specific patient subpopulation *c* ∈ *C*, where *C* represents all patient groups (e.g., labels in a classification problem or response population in survival analysis), the aggregated mean cosine similarity of a functional group ζ*_c_* over the subpopulation *c* represents the effect of the functional group in the subpopulation. Therefore, mean cosine similarities close to 1 indicate that the functional group is characteristic of the subpopulation, whereas a mean cosine score close to 0 reflects high orthogonality, indicating that the functional group is not associated with the given subpopulation. Core functional groups are those that show a high cosine similarity across all subpopulations, whereas target functional groups are enriched on specific subpopulations.

Therefore, we ranked the functional groups by their mean cosine similarity on each subpopulation to describe their most informative functional groups.

#### Validation of mutational functional groups

The mutational data offer a direct alternative to validate the effect of functional groups from the input data. In particular, we can measure the impact of mutations in patient survival outcome by binarizing each functional group to either mutation or wild-type status. Formally, a functional group is defined as a collection of mutated genes, ζ*_k_* = {*g*_1_,…,*g_i_*,…,*g_Gk_*}, where *G_k_* represents the genes in the group *k*. Therefore, for a given patient *I*, we can measure whether the functional group *ζ*^-^ is mutated if at least one gene in *G_k_* is mutated as follows:

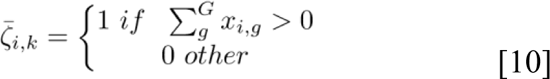

where *k* represents the *k*th functional group and *x_i_*_,*g*_ represents the gene value *g* for patient *i*. We then fit a univariate Cox PH model for each binary functional group, selected only those groups that are statistically significant (*P* < 0.05), and ranked them based on their HR. Statistically significant functional groups were tested on the complete training set and on a fully independent validation data set.

#### Functional group simplification of survival modeling

A subset of functional groups are strongly associated with patient survival. To maximize the signal from the functional groups, we trained a multivariate Cox PH regression model using the top 10 binarized functional groups *ζ*^-^ as input. This model uses a subset of input features (aggregated genes as functional groups), reducing the model’s complexity. To evaluate the performance of the model, we used 10 training and testing splits of 80% and 20%, respectively, over the Samstein et al.^25^ pan-cancer data set. To evaluate the model on independent data sets, we trained a Cox PH model using the entire Samstein et al.^25^ data set. The model outperformed TMB and achieved comparable performance to the clinical transformer model trained with all input features (genes). The multi–Cox PH model was also compared with a control model in which random functional groups ζ^rand^ were built by randomly selecting the same number of genes as the real functional groups ζ to train a multivariate Cox PH model. The multivariate Cox PH model was evaluated in the pan-cancer and cancer-specific settings across multiple studies and trials.

### Clinical transformer as generative model

To extract potential trajectories in the single-variable perturbation setting, we perturbed the input feature *f*_*i*_ by sampling the feature according to its distribution in the training population (i.e., divided the distribution of feature *f*_*i*_ across all patients into 10 percentiles and used its corresponding value as perturbation). This generated a new set of output embeddings along with their corresponding survival scores. On the other hand, to measure the impact of two feature interactions (pairwise interactions), we randomly sampled each pair of feature values from the training data while keeping all other features constant. We repeated this process 50 times for each feature pair and each patient. We were then able to identify the features and interactions with the strongest impact on the model’s output (survival score).

### Feature-feature interactions from the generative model for patient survival scores

Let *P* be the population of patients and let *F* be the set of features. For each patient *p* in *P*, we performed *F* single simulations by perturbing one feature at a time, and *F* × *F* paired simulations by perturbing two features at a time.

For each simulation, we recorded the patient’s maximum survival score. Let *M*_*p*_(*f*) denote the maximum survival score for patient *p* in the simulation where feature *f* is perturbed, and let *M*_*p*_(*f*, *g*) denote the maximum survival score for patient *p* in the simulation where features *f* and *g* are perturbed. For each feature *f*, we have a distribution of maximum survival scores, consisting of *M*_*p*_(*f*) for all patients in *P*. Similarly, for each feature pair (*f*, *g*), we have a distribution of maximum survival scores, consisting of *M*_*p*_(*f*, *g*) for all patients in *P*.

We computed the Mann-Whitney nonparametric statistical test between the distribution of maximum survival scores for each feature and feature pair and the distribution of the survival scores from the original data (without perturbation). Let *P*_*val*_ (*f*) and *P*_*val*_(*f*, *g*) be the *P* values resulting from the statistical test for feature *f* and feature pair (*f*, *g*), respectively. We transformed *P*_*val*_(*f*) and *P*_*val*_(*f*, *g*) to the –log scale, denoted as −*log*(*P*_*val*_(*f*)) and – *log*(*P*_*val*_(*f*, *g*)), respectively. To evaluate the effect of an interaction being stronger than the individual counterparts, we used the following equation:

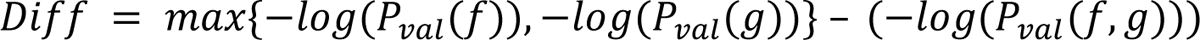

where *P*_*val*_(*f*)and *P*_*val*_ (*g*) are the –log of the *P* values for the features *f* and *g*, respectively, and *P*_*val*_(*f*, *g*) is the *P* value of the interaction between features *f* and *g*. If *Diff* is negative, it represents an aggregated value for the interaction between features *f* and *g*, indicating that the combined perturbation of both features is more significant than the individual perturbations. If *Diff* is positive, it indicates that the individual perturbations are enough to improve patient survival score.

### Analysis of variant and invariant populations

A feature or set of features *F* is perturbed *n* times, generating a conditional output patient embedding *P*^[*CLS*]^(*M*|*F* = *f*_*i*_), as well as a conditional survival score β(*M*|*F* = *f*_*i*_), where *M* represents the trained clinical transformer model and *f*_*i*_ a given perturbation *i* < *n* in the feature *f*. We can define the change in survival scores for a given patient *j* by taking the difference between the maximum and minimum of the perturbed survival scores β_j_^*f*^ = {β_j1_^*f*^, …, β_jn_^*f*^ } over the feature (or set of features). The Δβ_j_^*f*^ = max{β_j_^*f*^} − min{β_j_^*f*^} describes the impact of the perturbation of the feature *f* in the patient survival score, indicating the sensitivity of the patient *j* to a given perturbation *f*. We can then define two populations based on the median of the distribution of all patients Δβ_j_^*f*^: a variant population defining patients with a Δβ_j_^*f*^score higher than the median of the Δβ_j_^*f*^ distribution, and an invariant population defining patients with a Δβ_j_^*f*^below the median of the Δβ_j_^*f*^distribution. In the perturbation analysis for the Chowell et al.^3^ and Bagaev et al.^56^ data sets, we defined the variant/invariant populations exclusively on the poor and low-mid survivors in order to identify patients that under a perturbation show a transition that reflects improved survival.

## Data Availability

All data produced in the present work are contained in the manuscript

## Acknowledgments

All authors are employees of AstraZeneca and may hold stock ownership, options, or interests in the company.

