## supplementary material for "Pretrained transformers applied to clinical studies improve predictions of treatment efficacy and associated biomarkers"

### Supplementary Information

### A1 Clinical transformer training on the Chowell et al. data set for comparison with Chowell's model

To evaluate the clinical transformer' performance in the Chowell et al.[1] data set, we used the same training/testing splits provided in their study. We pretrained a model using all the data in the training split for 20,000 epochs and stored all 20,000 model weights. We fine-tuned a clinical transformer specialized for predicting patient overall survival using a training split in which 90% of the training data was used to train the model and 10% was used for validation. To identify the best pretrained model for this task, we selected several pretrained snapshot models at 100, 500, 1000, 5000, 15,000, and 20,000 pretraining epochs. For each pretrained model we then fine-tuned a survival model. We selected the 20,000 pretrained model snapshot for transfer learning because it provided the best performance in the validation set. The best epoch was selected based on the average concordance index (C-index) across the 10 runs in the validation set. Once the best pretrained weights (20,000) and epoch (195) were defined from the validation split, we trained a new model using 100% of the training split for 195 epochs. We evaluated this model in the 20% test set and compared it against tumor mutation burden (TMB) and the random forest model from Chowell et al. study.

### A2 Evaluation of clinical transformer trained using the Chowell et al. data set in the training/testing split framework and in the MYSTIC trial as independent data set

In this experiment, we merged the training and testing data from Chowell et al. into one single data set and evaluated the performance of the clinical transformer against other survival techniques with and without pretraining, repeating the process 10 times. First, the data were divided into 80% for training and 20% for testing samples (10 times). Second, we pretrained the clinical transformer model using the entire data set during 30,000 epochs. Third, we trained 10 clinical transformer models using the Chowell et al. training splits. Each fold was trained independently with and without the 30,000-pretraining snapshot. We empirically selected the best survival model epoch based on the mean C-index across 5% of the testing data, which was contrasted against the complete test set over the 10 splits (511 and 300 for the without and with pretraining models, respectively; MSI_SCORE and FCNA were unavailable in the MYSTIC trial [NCT02453282]). As the clinical transformer can handle missing features at inference, these two variables were not used in predicting patient survival scores.

The results showed that the model without pretraining achieved a C-index of 0.714, as compared with the model with pretraining, which had a C-index of 0.720; the random forest model, with a C-index of 0.714; Cox proportional hazards (PH), with a C-index of 0.709; and TMB, with a C-index of 0.55 (Table ST2.1).

We evaluated the performance of the clinical transformer using the Chowell et al. MYSTIC trial data by matching the features available on the trial. In total we identified a cohort of 150 patients (74 treated with PD-L1 and 76 with PD-L1 + CTLA-4) with a complete feature set (from 325 patients with available tissue TMB, only 150 had germline human leukocyte antigen [HLA] typing [HLA evolutionary divergence, HED]), except for MSI_SCORE and FCNA, which were unavailable in the clinical trial data. The 10 clinical transformer models trained on the Chowell et al. data were evaluated on MYSTIC data. The model with pretraining achieved a C-index of 0.643, whereas the clinical transformer without pretraining obtained a C-index of 0.616 and TMB on MYSTIC data showed a C-index of 0.608. Random forest and the Cox PH model were not evaluated on the MYSTIC data, as those models cannot handle missing data. For patient stratification (Supplementary Fig. SF2.1), we extracted the median cutoffs from the training splits for the different methods (the direct and gradual learning as well as for the TMB score; (Supplementary Table ST2.1, Supplementary Fig. SF2.1).

**Supplementary Table ST2.1.** Performance of the clinical transformer model trained on the Chowell et al. data set and evaluated on the MYSTIC trial

| **Modeling framework** | | **Chowell et al. 2021** | **MYSTIC (validation)** |
|---|---|---|---|
| Clinical transformer | Direct learning | 0.714 ± 0.01 | 0.616 ± 0.004 |
| | Gradual learning | **0.720 ± 0.01** | **0.643 ± 0.004** |
| Linear modeling | Cox PH regression | 0.709 ± 0.01 | — |
| Nonlinear modeling | Random survival forest | 0.714 ± 0.01 | — |
| Biomarkers | TMB | 0.550 ± 0.02 | 0.608 ± 0.000 |

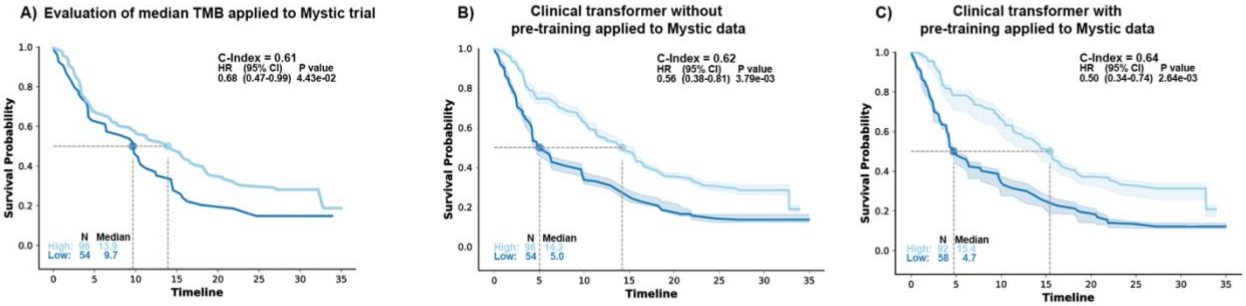


**Supplementary Fig. SF2.1.** Patient stratification using **a**, TMB; **b**, direct; and **c**, gradual learning in the test sets (10 repetitions). Patient population was stratified using the median cutoff from

the training splits. Solid line indicates the averaged KM curve across the 10 training repetitions and the areas represent the variability across the 20 testing splits.

### A3 Samstein et al. data set training/testing framework

We pretrained the clinical transformer using all patient population from GENIE v11 using all available clinical and molecular data during 20,000 iterations.

We trained a clinical transformer model to predict patient overall survival in the Samstein et al. data set, using 10 splits of 80% training and 20% testing. The baseline model corresponds to the model trained to predict survival without GENIE pretraining, and the E20000 model refers to the clinical transformer model trained on the Samstein et al. data, using the GENIE pretrained weights at the 20,000 snapshot.

Best epoch was empirically selected by looking at a small 5% proportion of the testing set in which 20% of the features were randomly shuffled to increase variability. We also evaluated the model's performance in the full testing set, using the average performance C-index across the 10 test sets. We did not find any significant difference in the testing at 5% and 100% evaluations. Therefore, we selected the models at the 25-training epoch (no difference in performance between epoch 23 to 30) for the pretrained model and the epoch 85 for the baseline model. Note that the same best epoch range (<25) was observed in an independent run using the Memorial Sloan Kettering Multimodal Integration of Data set (80/20% data splits) with the GENIE pretrained at the 20,000 snapshot. Independently, these two runs confirmed that the best model using pretraining lay approximately at epoch 25.

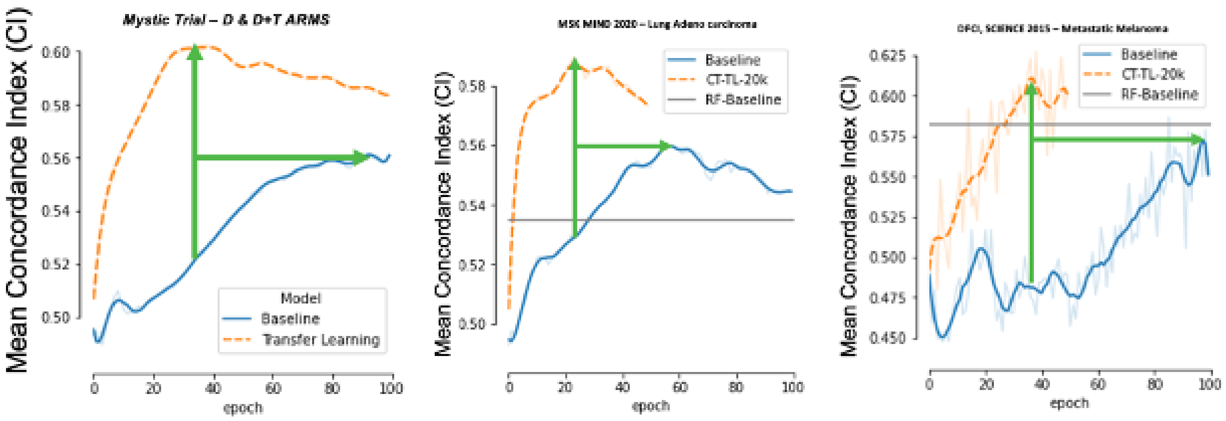


**Supplementary Fig. SF3.1.** Positive effect of transfer learning from pretrained model using GENIE to other small data sets. Models pretrained with GENIE data set achieved a peak performance in a smaller number of epochs compared with baseline clinical transformer trained models.

**Supplementary Table ST3.1.** Impact of GENIE transfer learning

| Learning type | Samstein et al.[2] | DFCI melanoma | MYSTIC trial | MSK MIND |
|---|---|---|---|---|
| **Concordance index** | | | | |
| Direct learning | 0.627 ± 0.02 | 0.587 ± 0.10 | 0.561 ± 0.05 | 0.560 ± 0.02 |
| Transfer learning | 0.649 ± 0.02 | 0.628 ± 0.07 | 0.602 ± 0.05 | 0.590 ± 0.03 |
| **Training epochs** | | | | |
| Direct learning | 86 | 85 | 95 | 57 |
| Transfer learning | 27 | 38 | 38 | 23 |
| Average reduction (%) | | | | 39.1130271 |

The GENIE pretrained model was used to fine-tune survival time and event across four data sets. **Top:** Concordance index on best model (averaged over 10 testing splits) for both direct and gradual learning. **Bottom**: Number of epochs (or iterations) the model needs to achieve peak performance.

### A4 Transfer learning: pretraining from the Chowell et al. data set and transfer to MYSTIC

We evaluated the added value of the transfer learning using the Chowell et al. data set over the MYSTIC data set. First, we pretrained a model using all the data from the Chowell et al. data set (train + test splits) and used the model snapshot at epoch 30,000. The pretrained model weights were transferred to a specialized model to predict patient overall survival using the MYSTIC data. For survival analysis, the MYSTIC data were divided into 10 training and testing splits, and 10 models were trained with and without pretraining. We selected the best model based on the averaged C-index of the 10 models in the test set. A random survival forest, a Cox PH model, and TMB were used for comparison (see Table 2 in the main article).

### A5 Transfer learning with GENIE to the Samstein et al. data set using only mutational data

We trained a clinical transformer model using only the mutational data from the Samstein et al. data set (469 gene mutations) without any other clinical or aggregated feature (e.g., race, age, TMB). This experiment was conducted to test the ability of the clinical transformer to identify molecular features associated with patient survival while being unbiased to any aggregated feature such as TMB. We used the pretrained GENIE data set at 20,000 epochs and trained the survival model for 10 repetitions, using splits of 80% training and 20% testing. The best model epoch was defined at 25 and was empirically selected as the average of the 10 model's performance in the test set. We evaluated the performance of the model using C-index. Note that the model trained only with mutational data (20,000 epochs, E020000) underperformed compared with the model that included clinical data (20,000 epochs, E020000B) but still outperformed the baseline without pretraining. For model interpretability, we computed all pairs cosine-similarities over the 10 test sets for consistency and extracted 50 functional groups by using a hierarchical clustering algorithm (Fig SF5.1). These functional groups represent the molecular associations within the data and their impact on survival.

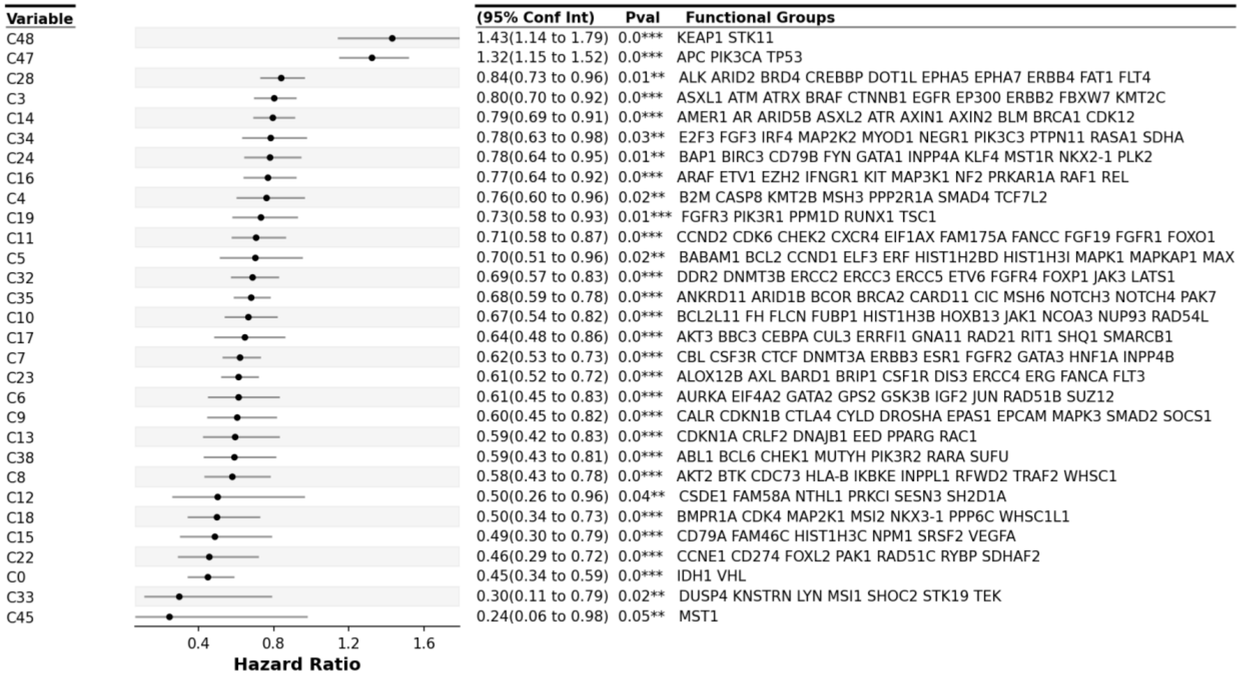


**Supplementary Fig. SF5.1.** Forest plot of significant ($P < 0.05$) functional groups.

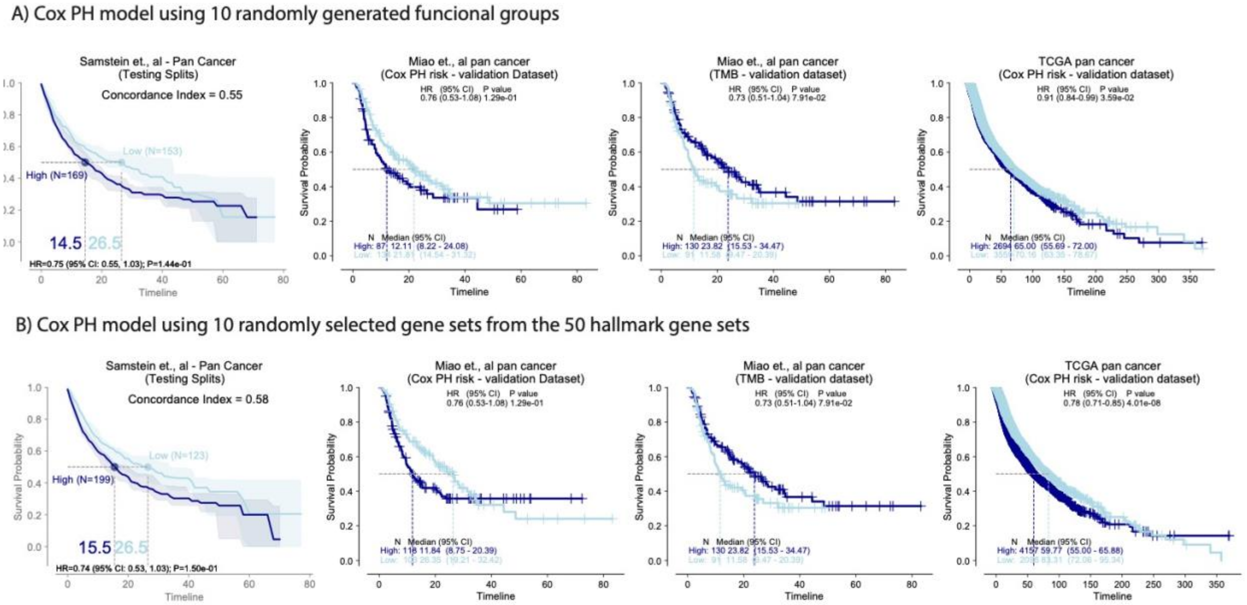


**Supplementary Fig. SF5.2.** Integration of functional groups to predict patient survival with immuno-oncology (IO) treatment in the testing splits (discovery data set) as well as on Miao et al.[3] pan-cancer IO-treated patients and treatment-naive pan-cancer data sets in The Cancer Genome Atlas (TCGA). **a**, Evaluation of randomly generated functional groups of the same size and number of genes as the top 10 groups. **b**, Evaluation of 10 randomly selected hallmark gene sets from the complete 50 hallmark sets and subsetting it to the MSK panel genes.

### A6 Identification of key functional groups associated with survival outcomes

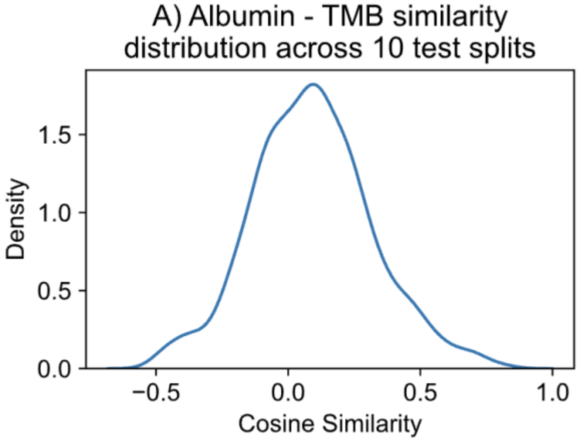


**Fig. SF6.1.** Albumin, TMB cosine similarity score across the test populations. Cosine similarity distribution.

**Figure ST6.1**: Pairwise cosine similarity between all feature embeddings across 10 test splits.

| source | target | cosine | cluster | Paper Cluster name |
|---|---|---|---|---|
| HED | Platelets | 0.24585439 | 0 | 2 |
| NLR | Platelets | 0.35590437 | 0 | 2 |
| Age | Platelets | 0.36766882 | 0 | 2 |
| Age | NLR | 0.38048447 | 0 | 2 |
| Albumin | HED | 0.38677867 | 0 | 2 |
| HED | NLR | 0.41395453 | 0 | 2 |
| BMI | HED | 0.42820412 | 0 | 2 |
| Age | HED | 0.44881286 | 0 | 2 |
| HGB | Platelets | 0.44995748 | 0 | 2 |
| BMI | NLR | 0.45971574 | 0 | 2 |
| Age | Albumin | 0.46831537 | 0 | 2 |
| HGB | NLR | 0.4693503 | 0 | 2 |
| Albumin | BMI | 0.47173626 | 0 | 2 |
| Albumin | NLR | 0.49050834 | 0 | 2 |
| HED | HGB | 0.49444037 | 0 | 2 |
| Chemo_before_IO (1:Yes; 0:No) | FCNA | 0.53687534 | 3 | 3 |
| Age | HGB | 0.53949961 | 0 | 2 |
| Albumin | Platelets | 0.55654177 | 0 | 2 |
| BMI | HGB | 0.55801385 | 0 | 2 |
| Age | BMI | 0.59264339 | 0 | 2 |
| MSI_SCORE | TMB | 0.59387972 | 2 | 1 |
| BMI | Platelets | 0.60135009 | 0 | 2 |
| Albumin | HGB | 0.60693464 | 0 | 2 |
| Chemo_before_IO (1:Yes; 0:No) | Sex (1:Male; 0:Female) | 0.62280129 | 3 | 3 |
| FCNA | Sex (1:Male; 0:Female) | 0.66104961 | 3 | 3 |
| HLA_LOH | MSI_SCORE | 0.67535925 | 2 | 1 |
| HLA_LOH | TMB | 0.69603414 | 2 | 1 |
| Cancer_Type | Drug_class | 0.97210378 | 1 | 4 |
| Cancer_Type | Stage at IO start | 0.9750603 | 1 | 4 |
| Drug_class | Stage at IO start | 0.98040073 | 1 | 4 |

### A7 Variant/invariant populations in the Chowell et al. data set

Probabilistic impact on patient survival is measured by the standard deviation of the distribution of all survival scores when one or two variables are perturbed. If a patient has a high standard deviation, it reflects changes in survival scores, whereas patients with standard deviation close to 0 indicate that the patient is not sensitive to perturbations in the given variable(s).

### A8 Perturbation of a T-cell gene expression signature identifies potential drivers of survival and resistance to Immune checkpoint inhibitor treatment

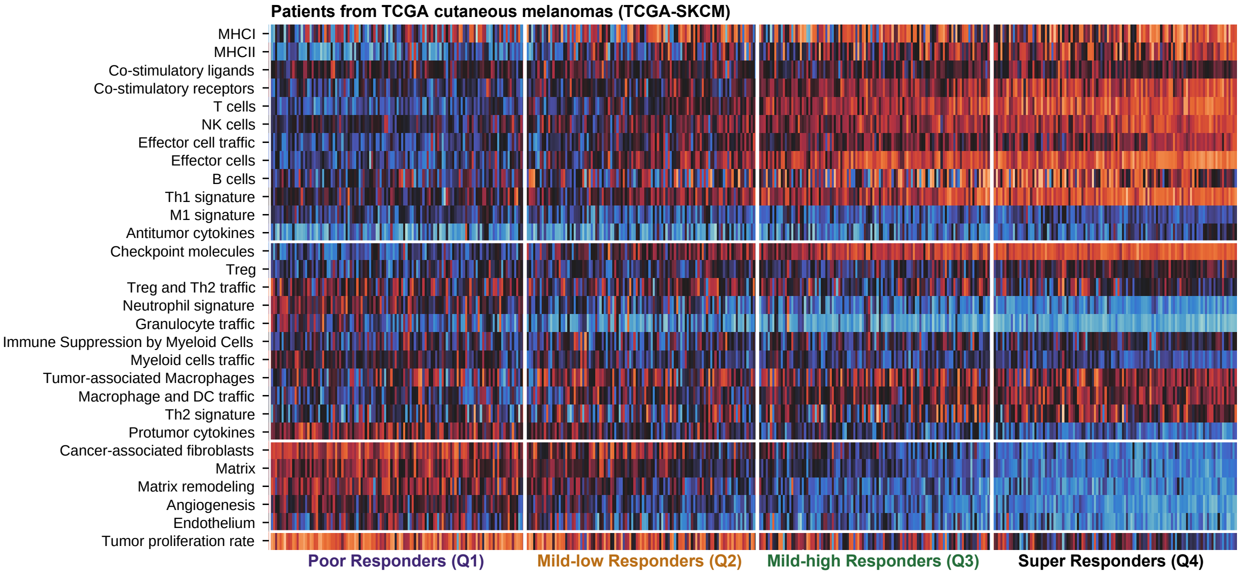


**Supplementary Fig. SF8.1.** Distribution of gene expression signatures across the four survival groups. Groups Q3 and Q4, with prolonged survival, demonstrated an increase in expression of signatures associated with productive antitumor immunity, such as major histocompatibility complex, T-cells, and T helper type 1 cell signaling. In contrast, groups Q1 and Q2, with reduced survival, demonstrate reduced expression of these signatures commensurate with an increase in signatures of neutrophils, protumor inflammatory signals, cancer-associated fibroblasts, and matrix remodeling.

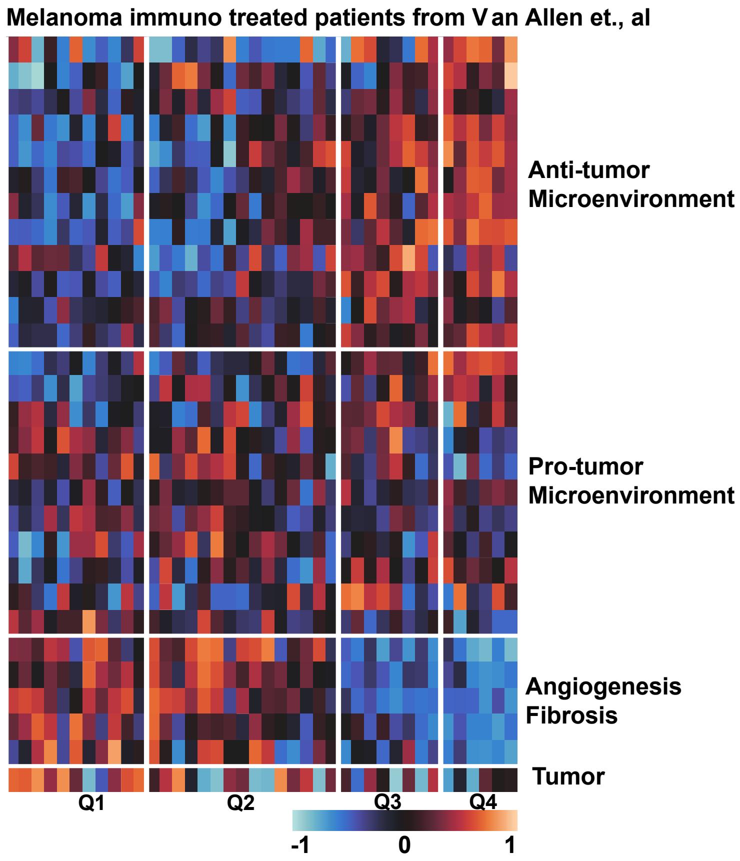


**Supplementary Fig. SF8.2.** Heat map of the tumor microenvironment signatures in the Van Allen et al. data set.[4]

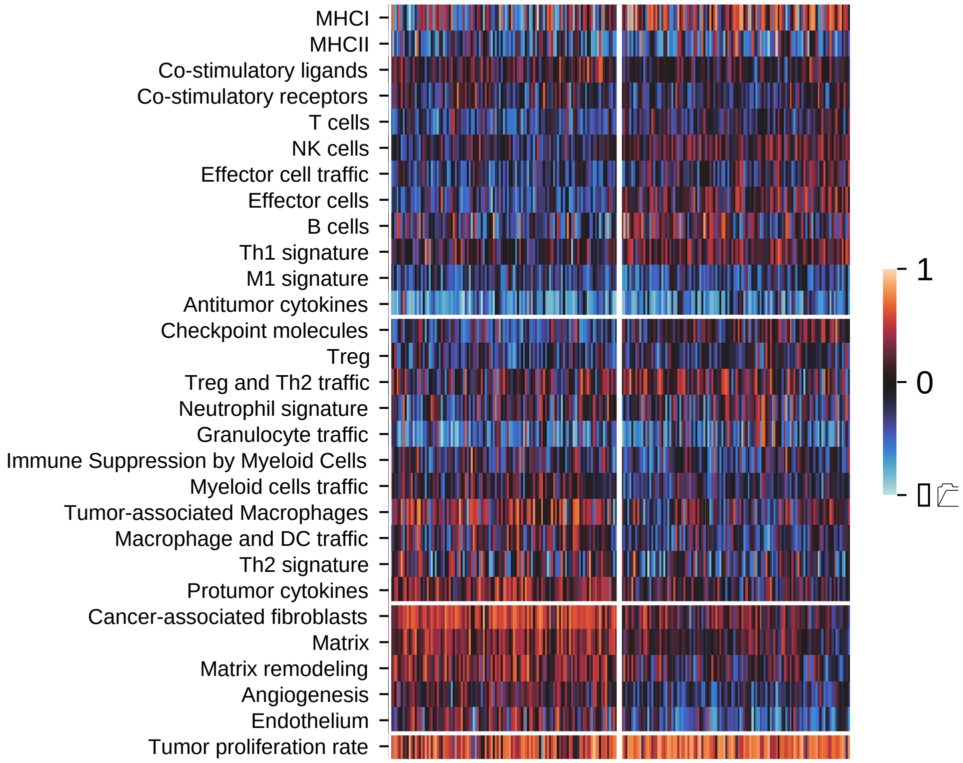


**Supplementary Fig. SF8.3.** Heat map of tumor microenvironment enrichment for the variant and invariant populations.

### Supplementary Table ST8.1

| feature | pval | Mean variant | Mean invariant | delta |
|---|---|---|---|---|
| Endothelium | 1.52062303234922E-15 | -0.313412674 | 0.112600806 | -0.426013479 |
| Cancer-associated fibroblasts | 4.06913332351783E-20 | 0.128167853 | 0.498169156 | -0.370001303 |
| Angiogenesis | 2.23255689268839E-15 | -0.200509689 | 0.120291558 | -0.320801246 |
| Matrix remodeling | 1.42213268012074E-10 | -0.014565958 | 0.28148602 | -0.296051978 |
| Matrix | 1.5429817283069E-14 | 0.057901648 | 0.331919433 | -0.274017785 |
| Protumor cytokines | 8.99938237234562E-11 | -0.000761741 | 0.269065383 | -0.269827124 |
| Tumor-associated Macrophages | 2.99159370163368E-06 | -0.015938159 | 0.221935134 | -0.237873292 |
| Macrophage and DC traffic | 5.58335783258359E-06 | -0.189935965 | 0.02157267 | -0.211508635 |
| Myeloid cells traffic | 2.53655426154648E-07 | -0.164258572 | 0.025172696 | -0.189431268 |
| Th2 signature | 0.005889151 | -0.160534751 | -0.010702139 | -0.149832612 |
| Immune Suppression by Myeloid Cells | 0.009670292 | -0.176916701 | -0.060527783 | -0.116388918 |
| Co-stimulatory ligands | 0.041212995 | 0.013129773 | 0.093677403 | -0.08054763 |
| Antitumor cytokines | 0.034964099 | -0.486369988 | -0.567646764 | 0.081276776 |
| Treg and Th2 traffic | 0.024231009 | 0.137531893 | 0.032257012 | 0.105274881 |
| Tumor proliferation rate | 1.81979544193861E-06 | 0.632009267 | 0.458450045 | 0.173559222 |
| B cells | 0.000349707 | 0.054694791 | -0.132986461 | 0.187681252 |
| Th1 signature | 5.72138985843501E-07 | 0.18682555 | -0.001916572 | 0.188742123 |
| T cells | 1.12293993693699E-07 | -0.081965987 | -0.280401733 | 0.198435746 |
| Effector cell traffic | 2.82314638046175E-06 | -0.044440517 | -0.247127222 | 0.202686705 |
| NK cells | 3.48706604111788E-12 | 0.150490246 | -0.115612173 | 0.266102419 |
| Checkpoint molecules | 3.97645311556161E-10 | 0.053729001 | -0.233654563 | 0.287383564 |
| MHCI | 7.6346147569635E-06 | 0.213198128 | -0.091958146 | 0.305156274 |
| Effector cells | 2.99686757789225E-16 | 0.130232828 | -0.271111321 | 0.401344149 |

### Supplementary Table ST8.2

| Population | hr | hr_lo | hr_hi | pval | N_High | N_Low | cohort | var |
|---|---|---|---|---|---|---|---|---|
| IO | 0.53141082 | 0.40298061 | 0.70077183 | 7.50E-06 | 227 | 227 | tcga | Effector cells |
| IO | 0.53032084 | 0.24673068 | 1.13986711 | 0.10423856 | 22 | 18 | allen | Effector cells |
| IO | 0.53338112 | 0.25117768 | 1.13264609 | 0.10188323 | 24 | 18 | liu | Effector cells |
| IO | 0.35451605 | 0.12128437 | 1.03625577 | 0.05810732 | 12 | 14 | riaz | Effector cells |
| IO | 0.85388965 | 0.64953366 | 1.12254003 | 0.25774627 | 227 | 227 | tcga | MHCI |
| IO | 0.76860871 | 0.3553994 | 1.66224068 | 0.50367427 | 17 | 23 | allen | MHCI |
| IO | 0.65615835 | 0.31400182 | 1.37115058 | 0.26248648 | 24 | 18 | liu | MHCI |
| IO | 0.49896671 | 0.18038488 | 1.38020314 | 0.18049481 | 12 | 14 | riaz | MHCI |
| IO | 0.47893362 | 0.36333878 | 0.63130453 | 1.75E-07 | 227 | 227 | tcga | Checkpoint molecules |
| IO | 0.53047203 | 0.2466364 | 1.14095314 | 0.10469667 | 20 | 20 | allen | Checkpoint molecules |
| IO | 0.59325921 | 0.28020074 | 1.25608698 | 0.1724949 | 27 | 15 | liu | Checkpoint molecules |
| IO | 0.95437163 | 0.35416572 | 2.57174863 | 0.92642876 | 11 | 15 | riaz | Checkpoint molecules |
| IO | 0.42085914 | 0.31750045 | 0.55786507 | 1.76E-09 | 227 | 227 | tcga | NK cells |
| IO | 0.43066657 | 0.19920376 | 0.93107526 | 0.03223313 | 22 | 18 | allen | NK cells |
| IO | 0.65372763 | 0.31520277 | 1.35582506 | 0.25342494 | 25 | 17 | liu | NK cells |
| IO | 0.51602203 | 0.17717994 | 1.5028718 | 0.22511322 | 11 | 15 | riaz | NK cells |
| IO | 1.75409344 | 1.33328858 | 2.30771031 | 5.94E-05 | 227 | 227 | tcga | Matrix remodeling |
| IO | 3.65286883 | 1.63044957 | 8.18390885 | 0.0016453 | 20 | 20 | allen | Matrix remodeling |
| IO | 1.46327324 | 0.66782985 | 3.20615884 | 0.34150812 | 15 | 27 | liu | Matrix remodeling |
| IO | 1.8890186 | 0.68270093 | 5.22687326 | 0.22061269 | 14 | 12 | riaz | Matrix remodeling |
| IO | 1.94989612 | 1.4747894 | 2.57805954 | 2.78E-06 | 227 | 227 | tcga | Angiogenesis |
| IO | 1.93459675 | 0.90157541 | 4.15124961 | 0.09026745 | 19 | 21 | allen | Angiogenesis |
| IO | 1.86582008 | 0.892888 | 3.89890401 | 0.09718239 | 19 | 23 | liu | Angiogenesis |
| IO | 1.80465743 | 0.62593523 | 5.20307577 | 0.27449724 | 15 | 11 | riaz | Angiogenesis |
| IO | 1.97472489 | 1.49685053 | 2.60516219 | 1.48E-06 | 227 | 227 | tcga | Cancer-associated fibroblasts |
| IO | 1.98349616 | 0.91511362 | 4.29920057 | 0.08270364 | 21 | 19 | allen | Cancer-associated fibroblasts |
| IO | 1.38564747 | 0.64212881 | 2.99008377 | 0.40588084 | 14 | 28 | liu | Cancer-associated fibroblasts |
| IO | 1.07221958 | 0.37089955 | 3.09963929 | 0.89755933 | 18 | 8 | riaz | Cancer-associated fibroblasts |
| IO | 1.87863847 | 1.4187695 | 2.48756582 | 1.07E-05 | 227 | 227 | tcga | Endothelium |
| IO | 1.01576509 | 0.46353656 | 2.22588421 | 0.96882738 | 16 | 24 | allen | Endothelium |
| IO | 0.87092976 | 0.41079463 | 1.84646682 | 0.71852204 | 18 | 24 | liu | Endothelium |
| IO | 1.9547557 | 0.70692985 | 5.40516122 | 0.19648771 | 14 | 12 | riaz | Endothelium |
